# Musculoskeletal Modeling and Movement Simulation for Structural Hip Disorder Research: A Scoping Review of Methods and Applications

**DOI:** 10.1101/2024.05.06.24306929

**Authors:** Margaret S. Harrington, Stefania D. F. Di Leo, Courtney A. Hlady, Timothy A. Burkhart

## Abstract

Musculoskeletal modeling is a powerful tool to quantify biomechanical factors typically not feasible to measure *in vivo,* such as hip contact forces and deep muscle activations. The purposes of this review were to summarize current modeling and simulation methods in structural hip disorder research and evaluate model validation practices and study reproducibility. MEDLINE and Web of Science were searched to identify literature relating to the use of musculoskeletal models to investigate structural hip disorders (i.e., involving a bony abnormality of the pelvis, femur, or both). Forty-seven articles were included for analysis. Studies either compared multiple modeling methods or applied a single modeling workflow to answer a research question. Overall, differences in outputs were shown between generic models scaled to participants’ anthropometrics and models with additional patient-specific geometry; however, generic models were most commonly used in application studies. The 11 studies that assessed model validation used qualitative approaches only. There was also wide variability and under-reporting of data collection, data processing, and modeling methods. Common assumptions made in musculoskeletal modeling during the model development, validation, and movement simulations were identified that are important to consider when evaluating the clinical applicability of modeling predictions in patients with structural hip disorders. Differences between generic and patient-specific model outputs exist; however, whether the patient-specific models are more accurate is still unknown. Increased transparency in reporting of data collection, signal processing, and modeling methods is needed to increase study reproducibility and allow for better assessment of modeling results.

## 1. Introduction

Structural disorders of the hip, such as femoroacetabular impingement syndrome (FAIS) and hip dysplasia, are associated with abnormal intra-articular loading of the hip joint, which can result in pain, decreased hip function, and increased risk of osteoarthritis (Husen et al., 2023; Song et al., 2019). Understanding the relationship between movement strategies, muscle forces, and hip contact forces (HCFs) can provide insight into the mechanisms underlying the etiology and treatment effectiveness of structural hip disorders. However, quantifying HCFs and deep hip muscle forces is generally not feasible to measure *in vivo*, given the invasive procedures required to collect this data (Hicks et al., 2015). Musculoskeletal (MSK) modeling has provided a method to quantify these internal biomechanical factors based on non-invasive, experimentally-measured external kinetics and kinematics (Damsgaard et al., 2006; Seth et al., 2018).

A key challenge of MSK modeling is the need to simplify the complexity of the MSK system to minimize computational costs while maintaining appropriate accuracy based on the research question (Hicks et al., 2015). This has led to wide variability in the modeling and simulation methods, which has created difficulties in making comparisons between studies. Furthermore, the variability in model validation criteria has created challenges in assessing the clinical applicability of modeling study results. Therefore, the purposes of this scoping review were to: i) summarize current modeling and simulation methods in structural hip disorder research; ii) describe model validation practices; and iii) highlight current issues related to study reproducibility.

## 2. Methods

### 2.1. Search strategy

The search strategy was developed with the support of a professional librarian. The electronic databases MEDLINE and Web of Science were searched to identify relevant studies. The complete search strategies for each database are presented in Appendix A. In summary, the search strategies included keywords related to three search concepts: i) MSK modeling (e.g., models and simulations); ii) hip (e.g., pelvis and femur); and iii) structural disorder (e.g., disorder, pathology, and abnormality). This search strategy was developed based on a set of relevant studies that met the inclusion criteria. The retrieval of an additional set of pre-identified articles that met the inclusion criteria was used to validate the search strategy. Studies were retrieved up to July 3^rd^, 2023.

### 2.2. Inclusion and exclusion criteria

Studies were included if they developed or analyzed an MSK model to investigate a structural hip disorder. For this scoping review, a structural hip disorder was defined as any bony abnormality of the femoral neck, femoral head, or acetabulum (Clohisy et al., 2005). MSK modeling was defined as a computational representation of the muscular system acting on a rigid, multibody skeletal structure (Hicks et al., 2015). Studies were excluded if i) anatomical hip joint structures were not modeled or analyzed (e.g., they only investigated the effect of a prosthesis); ii) the experimental data collection included the use of walking aids (e.g., crutches, braces); iii) a non-human population was investigated; iv) it was written in a language other than English; and v) it was published as a conference proceeding only.

### 2.3. Study selection

Study duplicates from the search results were removed in EndNote before uploading the titles and abstracts to an online screening platform (Covidence 2023, Melbourne, Australia). Two of three independent reviewers screened all titles and abstracts of records for inclusion and exclusion. Then, two of three independent reviewers screened the full-text reports for studies included in the title and abstract screening step. Conflicts were resolved by a third reviewer (TB) at both stages of the screening process.

### 2.4. Quality assessment

A previously developed checklist for biomechanics research was used to assess the included studies’ quality (Appendix B) (Moissenet et al., 2017). The articles were scored for each question based on no (zero points), limited (one point), or satisfactory (two points) information, and the overall score was calculated as the sum of points divided by the maximum potential score for applicable questions (Moissenet et al., 2017). A sample of three studies was evaluated by two authors (MH, TB), and disagreements were discussed and resolved between the two assessors to reach a consensus on the interpretation of the quality assessment. For this study, the checklist was modified to include the question: “Were relevant instrumentation specifications and signal processing techniques described?” In addition, the questions “Was the evaluation strategy appropriately justified?” and “Were the analytical methods clearly described?” were removed because the other questions and the analysis conducted for the scoping review expanded on these areas of the studies. The evaluation of each study conducted by one author (MH) is reported in Appendix B (Table B1).

### 2.5. Data Extraction

A data extraction form in Microsoft Excel (Version 16.70, Microsoft Corporation, Redmond, USA) was developed to extract the required information for analysis, including the authors, publication date, sample demographics, primary study objective, main results, MSK model characteristics (and any alterations if a generic model was used), experimental inputs, simulation outputs, and validation procedures. Two of the three authors (MH, SD, CH) completed data extraction for each article, and disagreements were resolved in a discussion meeting. A thematic analysis was conducted to identify key issues and themes related to the MSK modeling methods and applications (Kiger and Varpio, 2020). If the models were incompletely described, details regarding MSK model characteristics or alterations to generic models were retrieved from references or the OpenSim modeling software (Stanford University, Stanford, USA) documentation (https://simtk-confluence.stanford.edu:8443/display/OpenSim/OpenSim+Documentation).

## 3. Results

### 3.1. Search results and quality assessment

A total of 1525 articles were retrieved from the database searches and 47 were retained for analysis (Figure 1). Summaries of the study characteristics and population demographics are provided in Table 1 and 2, respectively. The hip disorders investigated in the included studies were osteoarthritis (n = 15), hip dysplasia (n = 11), cerebral palsy-associated femoral deformities (n = 8), FAIS (n = 9), and idiopathic femoral version deformities (n = 4) (Table 1). Gait was the most common movement simulated (n = 36). Ten studies included simulations of more demanding and hip-provocative tasks, such as double and single leg squats (n = 5), stair climbing (n = 4), isolated hip range of motion (n = 3), and rehabilitation exercises (i.e., hip-focused with lower-extremity and trunk strengthening and stretching) (n = 1).

**Fig. 1.**
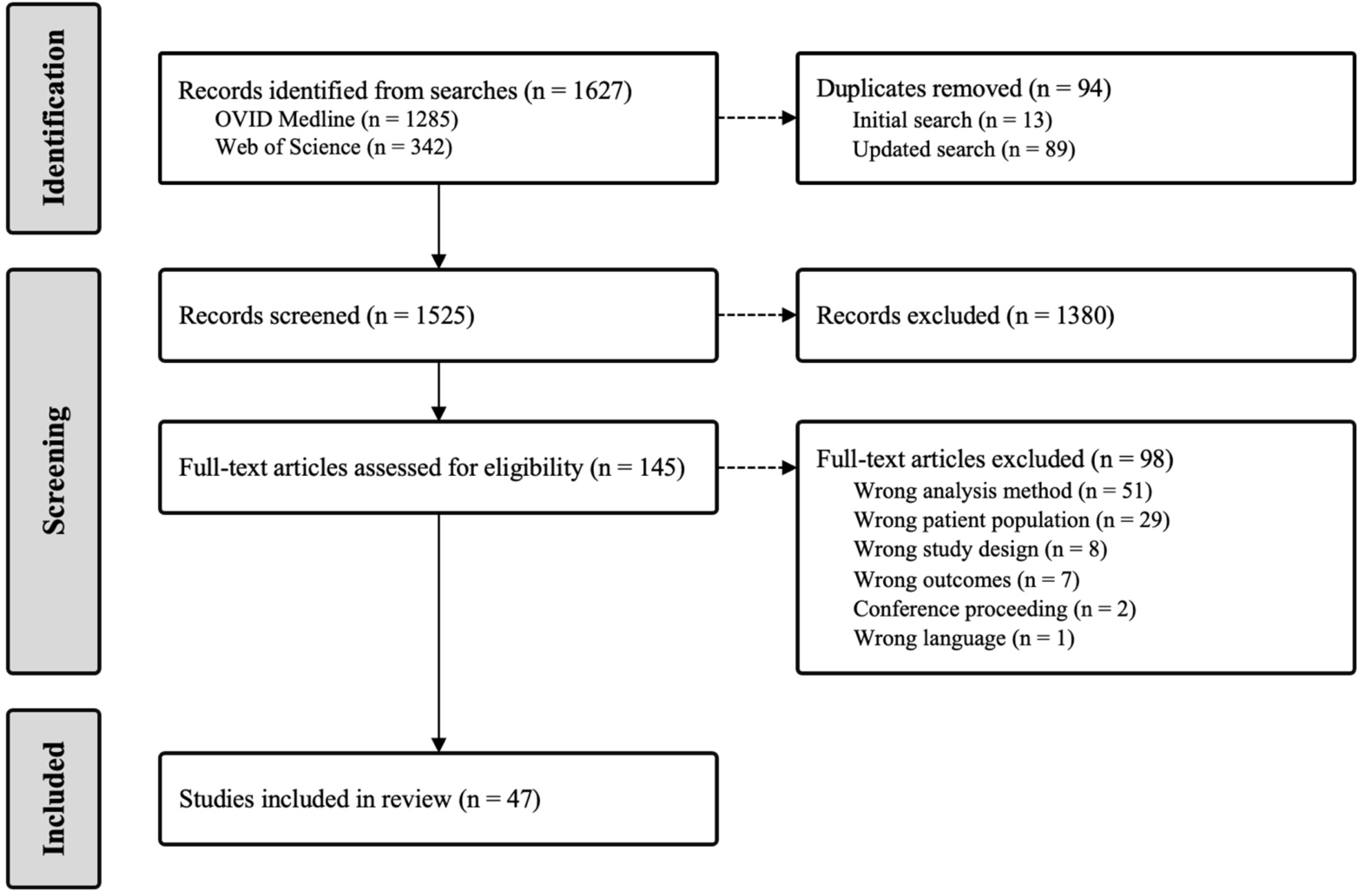
PRISMA Flow Chart.

**Table 1.**
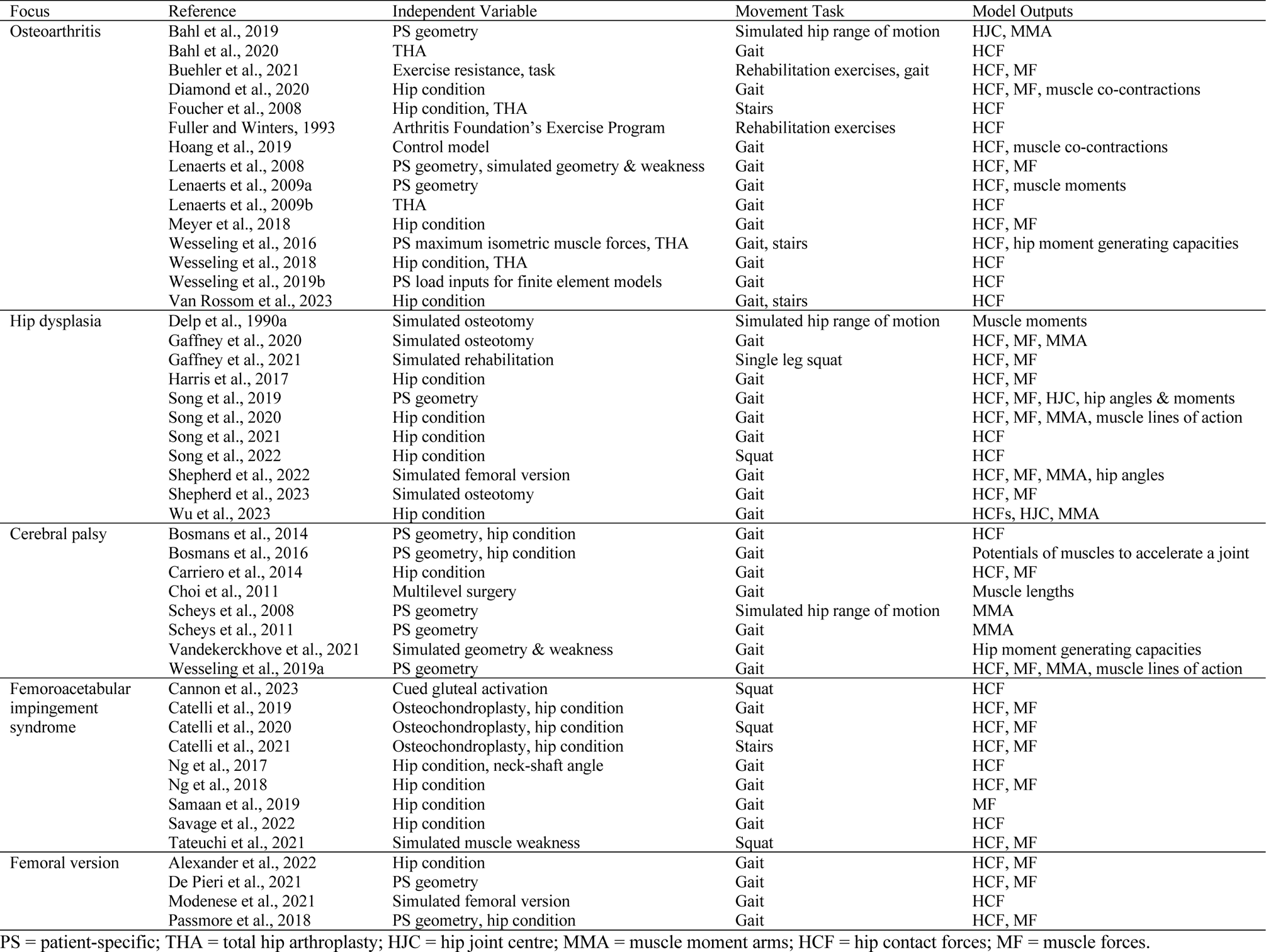
Overview of included studies’ independent variables, movement tasks simulated, and main model outputs of interest.

**Table 2.**
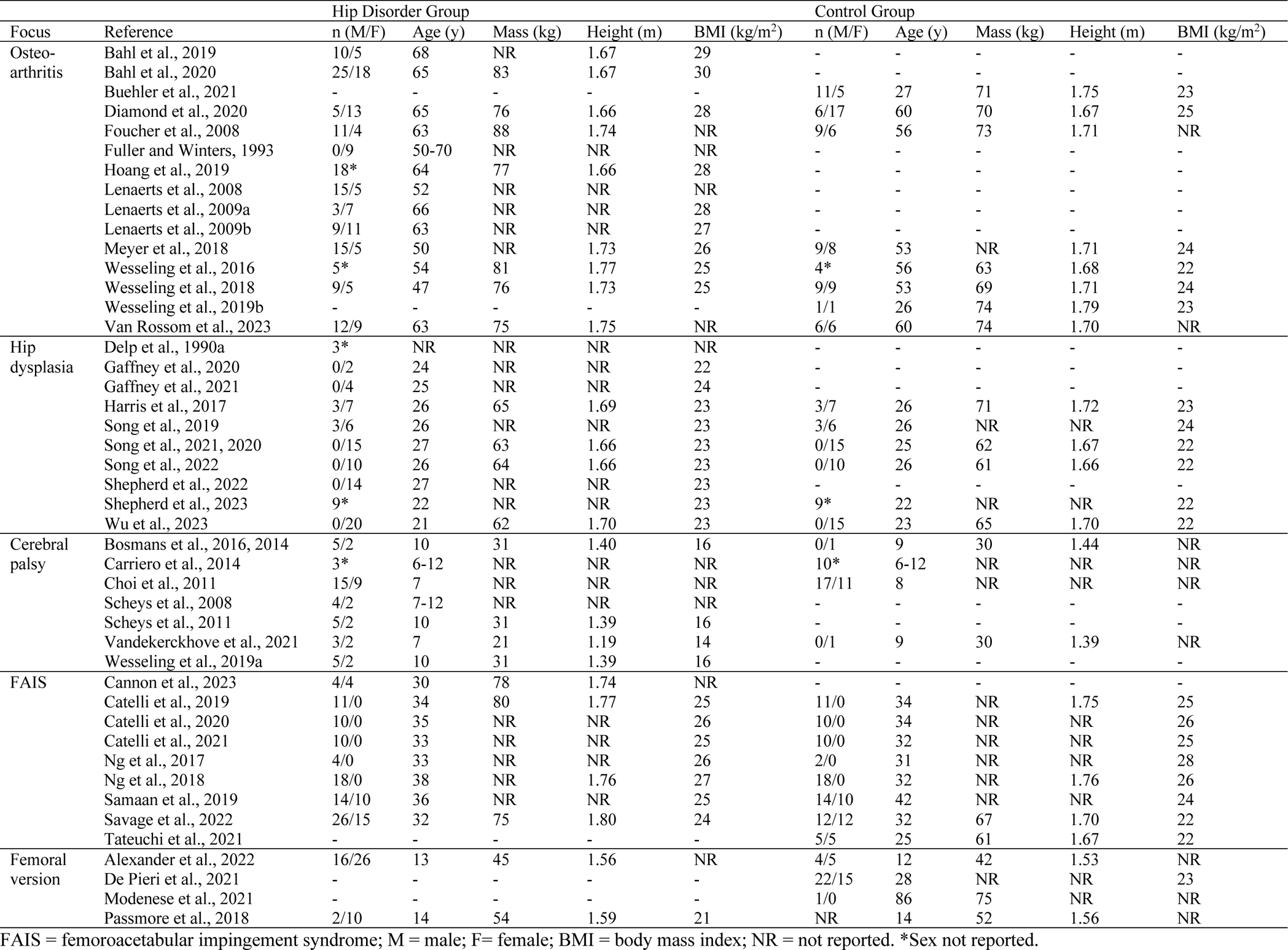
Overview of included studies’ sample size (n) and demographics reported as means, medians, or ranges.

The included studies primarily implemented MSK modeling to quantify HCFs (n = 39) and muscle forces or activations (n = 22) (Table 1). Main outcomes of interest reported also included parameters related to muscle paths and capacities (e.g., moment arm lengths, moment generating capacity), hip joint centre locations, joint angles and moments, and muscle co-contraction indices. Three studies took the MSK model outputs and used them as inputs into different types of models (e.g., finite element models) to quantify acetabular cartilage and labrum stresses (Ng et al., 2017) and acetabular contact pressure or edge loading (Cannon et al., 2023; Wesseling et al., 2019b).

Each study fell into one of two general categories: i) studies that compared different modeling and simulation methods in the context of a hip disorder (n = 16) (e.g., compared generic versus patient-specific geometry) which will be referred to as “methods studies” (Table 3); or ii) studies that applied a single modeling workflow to study a hip disorder (n = 31) which will be referred to as “application studies” (Table 4).

**Table 3.** Summary of studies that compared modeling and simulation methods.

| Reference | Comparisons | Additional Modeling Choices | Summary of Results and Conclusions |
| --- | --- | --- | --- |
| Bahl et al., 2019 | Four models:<br>1) Shape modeling, PC fit<br>2) Shape modeling, CT<br>3) SMS with functional HJC<br>4) SMS with regression HJC | *Model:* Gait2392 (Models 3 & 4)<br>*HJC:* CT-derived (Models 1 & 2)<br>*MTU:* Re-mapped generic MTU (Models 1 & 2) | *Results:* Mean HJC location error for statistical shape models 1 and 2 (11.4 mm, 6.6 mm) were significantly lower than scaled generic models 3 and 4 (36.9 mm, 31.2 mm). Mean RMSE were greatest for hip MMA lengths using generic models (16.15-16.71 mm) versus shape models (0.05-3.2 mm).<br>*Conclusion:* Shape modeling improves HJC location and MMA lengths. |
| Bosmans et al., 2014 | Four model and gait conditions:<br>1) Generic, normal gait<br>2) Generic, CP gait<br>3) MRI, normal gait<br>4) MRI, CP gait | *Model:* Gait2392 Iteration (Details not reported)<br>*Rigid Bodies:* SMS with global optimization<br>*HJC:* MRI-derived (Models 3 & 4)<br>*MTU:* MRI-based updated attachments & paths (Models 3 & 4)<br>*Control:* SO | *Results:* Using MRI models, CP gait patterns reduced HCFs and changed the orientation of the HCF vector more vertically and anteriorly compared to normal gait. More pronounced bony deformities were correlated with greater differences in HCF magnitude and orientations. Using generic models, the HCFs during CP gait were more similar to those shown during normal gait.<br>*Conclusion:* Femoral geometry influences HCFs. |
| Bosmans et al., 2016 | Three model and gait conditions:<br>1) Generic, CP gait<br>2) MRI, CP gait<br>3) MRI, normal gait | *Model:* Gait2392 Iteration (Details not reported)<br>*Rigid Bodies:* SMS with global optimization<br>*HJC:* MRI-derived (Models 2 & 3)<br>*MTU:* MRI-based updated attachments & paths (Models 2 & 3) | *Results:* The differences in muscles' potential to control joints between generic and MRI models were 398-894 ($^{\circ}/s^2$)/kg m, and 199-993 ($^{\circ}/s^2$)/kg m between gait conditions. MMA length changes were related to changes in potentials.<br>*Conclusion:* CP gait and femoral deformities have a concomitant effect on muscle control hip and knee motion, mainly in the sagittal plane. |
| De Pieri et al., 2021 | Two models:<br>1) Generic<br>2) Generic deformed to PS femoral version | *Model:* TLEM2<br>*Rigid Bodies:* SMS, radiograph-derived pelvic width scale factor<br>*Control:* SO | *Results:* There were significant differences in the HCFs between models across the complete gait cycle including more anteriorly directed forces. HCFs during mid to terminal stance were less proximally and medially directed.<br>*Conclusions:* PS femoral version may provide insight into joint damage risk. |
| Hoang et al., 2019 | Two control models:<br>1) SO<br>2) EMG-assisted | *Model:* Gait2392<br>*Rigid Bodies:* SMS<br>*HJC:* Regression<br>*MTU:* Optimized muscle fiber & tendon slack length | *Results:* Compared to SO, EMG-assisted model had better agreement with EMG (i.e., higher $R^2$ and lower RMSE) and higher levels of muscle co-contraction. EMG-assisted had higher HCFs than SO and *in vivo* HCFs.<br>*Conclusion:* EMG-assisted model solution can predict physiologically-plausible HCFs in a population with higher levels of muscle co-contraction. |
| Lenaerts et al., 2008 | 1) Sensitivity analysis of isolated changes in NL (40-80 mm) and NSA (110-150°)<br>2) Generic vs. deformed models (PS: NL & NSA) | *Model:* Gait2392 Iteration (7 segments, 16 DoF, 86 MTU)<br>*Rigid Bodies:* SMS<br>*Control:* SO | *Results:* Increasing NL increased the resultant HCF from 1.06 to 4.47 BW. NL predicted changes in all HCFs components' magnitudes and orientations in the frontal and sagittal planes. Increasing NSA increased gluteus medius and minimus activity. Increasing NL decreased gluteus medius activity.<br>*Conclusion:* NL alters HCFs, NSA only has minor effect on HCF. |
| Lenaerts et al., 2009a | Three models:<br>1) Generic<br>2) Generic deformed to PS femoral version, NL, & NSA<br>3) CT-derived pelvis | *Model:* Gait2392 Iteration (7 segments, 16 DoF, 86 MTU)<br>*Rigid Bodies:* SMS with global optimization<br>*HJC:* CT-derived (Model 3)<br>*Control:* SO | *Results:* Model 3 with CT-derived geometry and HJC location, shifted the HJC on average 30.3 mm anteriorly and 20.9 mm proximally. Model 3 had significantly lower mean peak hip flexion-extension moment compared to the generic Model 1 (0.415 vs. 0.704 Nm/kg) and increased mean frontal plane HCF inclination angle (32.59°) compared to Model 1 (16.35°) and 2 (16.05°).<br>*Conclusion:* Personalized geometry and HJC location affect HCFs. |
| Modenese et al., 2021 | Generic model deformed with femoral version between -2° to 40° | *Model:* Full Body<br>*Rigid Bodies:* Upper limbs removed, SMS<br>*MTU:* Attachments & paths deformed<br>*Control:* SO | *Results:* HCFs increased up to 17.9% with increasing femoral anteversion.<br>*Conclusion:* Femoral version substantially effects HCFs. |
| Passmore et al., 2018 | Two models:<br>1) Generic<br>2) PS femoral version, NSA, & tibial torsion | *Model:* Gait2392<br>*Rigid Bodies:* SMS<br>*HJC:* Radiograph-derived for patients<br>*MTU:* Attachments & paths deformed (Model 2)<br>*Control:* SO | *Results:* There were significant differences between models for hip muscle forces (RMSE = 0.05-0.18), and for HCFs (RMSE = 0.32-0.54).<br>*Conclusion:* Torsional deformities increase hip abductor force, with a corresponding increase in compressive HCFs. |

**Table 3** (*continued*)
| Reference | Comparisons | Additional Modeling Choices | Summary of Results and Conclusions |
| --- | --- | --- | --- |
| Scheys et al., 2008 | Two models:<br>1) Generic<br>2) MRI-derived geometry | *Model:* Gait2392 Iteration (7 segments, 16 DoF, 86 MTU)<br>*Rigid Bodies:* SMS with global optimization<br>*HJC:* MRI-derived (Model 2)<br>*MTU:* MRI-based updated attachments (Model 2) | *Results:* The mean difference in MMA lengths between models for hip sagittal, formal, and transverse plane muscles were 12.5%, 30.1%, and -96.5%.<br>*Conclusion:* Compared to MRI models, generic models overestimated MMA lengths for hip flexion, extension, abduction, adduction, and external rotation, and underestimate for hip internal rotation. |
| Scheys et al., 2011 | Three models:<br>1) Generic<br>2) Generic deformed to PS femoral version, NL, & NSA<br>3) MRI-derived geometry | *Model:* Gait2392 Iteration (Details not reported)<br>*Rigid Bodies:* SMS with global optimization<br>*HJC:* MRI-derived (Model 3)<br>*MTU:* Attachments & paths (Model 2), MRI-based updated attachments & paths (Model 3) | *Results:* Compared to the MRI model, the generic model on average overestimated the MMA lengths of the hip flexors (19.9%), extensors (24.7%), abductors (24.4%), and adductors (8.0%). Compared to the generic model, the deformed model only notably affected the MMA lengths for gluteus maximus.<br>*Conclusion:* Large differences in MMA lengths were shown between the generic and MRI models, which were not uniformly reduced by the deformed model. |
| Shepherd et al., 2022 | Deformed models of hip dysplasia patients with femoral version angles between -5° to 35° | *Model:* LaiArnold2017 Iteration (22 segments, 37 DoF, 92 MTU)<br>*Rigid Bodies:* SMS, MRI-derived pelvis/femurs<br>*HJC:* MRI-derived<br>*MTU:* MRI-based updated attachments & paths<br>*Control:* computed muscle control | *Results:* Increasing femoral anteversion resulted in the highest change of mean resultant HCFs in late stance (0.48 BW) and increased hip flexor and abductor muscle forces. HCFs were lowered by 0.32 BW on average with relative retroversion in late stance.<br>*Conclusions:* Increased anteversion is the strongest influence on HCFs. |
| Song et al., 2019 | Three models:<br>1) Generic<br>2) Generic with three CT-derived pelvis scale factors<br>3) CT-derived geometry | *Model:* 2396Hip<br>*Rigid Bodies:* SMS (one scale factor per segment)<br>*HJC:* CT-derived<br>*MTU:* MRI-based updated attachments & paths<br>*Control:* SO | *Results:* Mean resultant HCFs were significantly higher for CT-derived pelvis geometry (5.47 BW) versus the generic Model 1 (4.18 BW) and Model 2 (4.15 BW). The CT model showed significantly higher muscle forces compared to the generic models for hip flexors, extensors, internal rotators, and external rotators.<br>*Conclusions:* Geometry, HJC, and muscle paths affect HCFs and muscle forces. |
| Vandekerckhove et al., 2021 | 6400 models with simulated hip muscle weakness (0-75%), femoral version (20-60°) & NSA (120-160°). Simulated normal & CP gait patterns. | *Model:* Gait2392 Iteration (14 segments, 19 DoF, 88 MTU)<br>*MTU:* MIMF scaled to participants' mass<br>*Control:* SO | The capability gap (CG) in hip moment generating capacity increased by: 0.005-0.080 Nm/kg per 10% hip abductor weakness increase; 0.011-0.211 Nm/kg per 10° femoral anteversion increase; and with 0.011-0.163 Nm/kg per 10° NSA increase.<br>*Conclusion:* Increases in hip abductor weakness, femoral anteversion, and NSA predicted decreases in hip moment generating capacity at the hip. |
| Wesseling et al., 2016 | Two MIMF scaling methods:<br>1) static: dynamometer<br>2) functional: strength required to generate joint moments during movements | *Model:* Gait2392 Iteration (14 segments, 19 DoF, 88 MTU)<br>*Rigid Bodies:* SMS, MRI-derived geometry<br>*HJC:* based on femoral bone structures<br>*MTU:* MRI-based updated attachments & paths<br>*Control:* SO | *Results:* Functional scale factors were higher than the static for abductors (median = 1.10 vs. 0.54) and flexors (0.94 vs. 0.41). Statically scaled models lacked sufficient strength to generate joint moments required for the movements. HCFs were similar between functionally scaled and unscaled models.<br>*Conclusion:* Scaling model muscle forces is not required when quantifying HCFs if model is sufficiently strong. |
| Wesseling et al., 2019a | Three models:<br>1) Generic<br>2) Deformed pelvis, femurs, & proximal tibia<br>3) MRI-derived pelvis, femurs, & tibias | *Model:* Gait2392 Iteration (14 segments, 19 DoF, 88 MTU)<br>*Rigid Bodies:* SMS<br>*HJC:* MRI-derived<br>*MTU:* Attachments & paths deformed (Model 2), MRI-based updated attachments (Model 3)<br>*Control:* SO | *Results:* There were higher differences between the generic and MRI model than between the deformed and MRI model for muscle position (2.19 vs. 1.73 cm), MMA length (3.34 vs. 2.13 cm), and muscle forces (6.53 vs. 3.08 N/kg). The generic model had lower peak HCFs than the MRI model (3.15 vs. 4.99 BW).<br>*Conclusion:* The deformed model is more similar to the MRI model than the generic but cannot yet serve as a replacement for the MRI model. |

PC = principal components; CT = computed tomography; SMS = surface marker scaling; HJC = hip joint centre; CP = cerebral palsy; MRI = magnetic resonance imaging; PS = patient-specific; SO = static optimization; EMG = electromyography; NL = neck length; NSA = neck-shaft angle; MIMF = maximum isometric muscle force; MTU = musculotendon units; DoF = degrees of freedom; RMSE = root mean squared error; MMA = muscle moment arms; HCF = hip contact force.

**Table 4.** Modeling choices for the included studies that applied a generic model to answer a research question.

| Model | Reference | Rigid Bodies | HJC | Musculotendon Units | Control |
| --- | --- | --- | --- | --- | --- |
| Lower Extremity Model (7 segments, 7 DoF, 43 MTU) | Delp et al., 1990a | NR | NR | NR | N/A |
| Gait2392 (14 segments, 23 DoF, 92 MTU) | Bahl et al., 2020 | Statistical shape modeling | CT-derived | NR | SO |
|  | Buehler et al., 2021 | SMS | Regression | MIMF scaled to participants' mass | SO |
|  | Diamond et al., 2020 | SMS | NR | Optimized muscle fiber & tendon slack length | EMG-assisted |
|  | Ng et al., 2018 | SMS, CT-derived pelvis scale factors | NR | MIMF scaled to participants' mass & height | SO |
| Gait2392 Iteration (14 segments, 19 DoF, 88 MTU) | Meyer et al., 2018 | SMS | NR | NR | SO |
|  | Wesseling et al., 2018 | SMS | NR | Muscle attachments, fiber & tendon slack lengths scaled with rigid body scale factors | SO |
| Gait2392 Iteration (7 segments, 16 DoF, 86 MTU) | Carriero et al., 2014 | SMS | NR | NR | SO |
|  | Lenaerts et al., 2009b | SMS | NR | NR | SO |
| Gait2392 Iteration (8 segments, 19 DoF, 92 MTU) | Samaan et al., 2019 | SMS | NR | NR | CMC |
| Gait2392 Iteration (3 DoF knee) | Van Rossom et al., 2023 | SMS | NR | NR | SO |
| Gait2392 Iteration (34 MTU) | Savage et al., 2022 | SMS | Regression | Optimized muscle fiber & tendon slack length<br>MIMF scaled to participants' mass & height | EMG-assisted |
| Gait2392 Iteration (version not specified) | Choi et al., 2011 | NR | NR | NR | N/A |
|  | Foucher et al., 2008 | NR | NR | NR | Constraints |
|  | Wesseling et al., 2019b | SMS, MRI-derived pelvis, femurs, tibias, & patellae | MRI-derived | Hip & knee muscle attachments updated based on MRI & bony landmarks | SO |
| Full Body Running (12 segments, 29 DoF, 92 MTU) | Ng et al., 2017 | NR | NR | NR | SO |
| Fully Body (22 segments, 37 DoF, 80 MTU, 17 ITA) | Catelli et al., 2019 | SMS, 10x weighting for markers verified with CT | NR | NR | SO |
| LaiArnold2017 (22 segments, 37 DoF, 80 MTU, 17 ITA) | Gaffney et al., 2020 | SMS, MRI-derived pelvis & femurs | MRI-derived | Hip & knee muscle attachments updated based on MRI & bony landmarks | SO |
| Full Body Squat (22 segments, 37 DoF, 80 MTU, 17 ITA) | Catelli et al., 2021, 2020 | SMS, 10x weight CT verified markers verified with CT | NR | NR | SO |
|  | Gaffney et al., 2021 | SMS, MRI-derived pelvis & femurs | MRI-derived | Hip & knee muscle attachments updated based on MRI, MIMF scaled to torque data | CMC |
| 2396Hip (14 segments, 23 DoF, 96 MTU) | Harris et al., 2017 | SMS, CT-derived pelvis | CT-derived | Hip muscle attachments & paths updated based on MRI & bony landmarks | SO |
|  | Shepherd et al., 2023 | SMS, MRI-derived pelvis & femurs | MRI-derived | Hip muscle attachments & paths updated based on MRI & bony landmarks | CMC |
| 2398Hip (14 segments, 23 DoF, 98 MTU) | Song et al., 2022, 2021, 2020; Wu et al., 2023 | SMS, MRI-derived pelvis & femurs | MRI-derived | Hip muscle attachments & paths updated based on MRI & bony landmarks | SO |
| TLEM2 (11 segments, 21 DoF, 110 MTU) | Alexander et al., 2022 | SMS, personalized femoral version | NR | NR | SO |
|  | Tateuchi et al., 2021 | SMS | NR | Simulated hip muscle weakness | SO |
| Custom Models | Fuller and Winters, 1993 | NR | NR | NR | Heuristic |
|  | Cannon et al., 2023 | NR | NR | NR | EMG-driven |

DoF = degrees of freedom; MTU = musculotendon units; ITA = ideal torque actuators; NR = not reported; SMS = surface marker scaling; CT = computed tomography; MRI = magnetic resonance imaging; HJC = hip joint centre; MIMF = maximum isometric muscle force; SO = static optimization; CMC = computed muscle control.

### 3.2. Generic model development

Generic baseline models developed for the OpenSim or AnyBody Modeling System (AnyBody Technology, Aalborg, Denmark) software were used in 45 of the 47 studies (Tables 3 and 4). The remaining two studies used a custom algorithm (Fuller and Winters, 1993) and an electromyography (EMG)-driven model programmed in MATLAB (Cannon et al., 2023). An iteration of the OpenSim Gait2392 model was most commonly used (n = 26, Tables 3 and 4). An issue identified was that several studies indicated they used a version of the Gait2392 model or the original Lower Extremity Model developed by Delp et al. (1990b), but did not specify which iteration. Fifteen studies used a different OpenSim generic model, and three used the generic Anybody Twente Lower Extremity Model Version 2 (TLEM2).

Thirty-three of the studies used generic models with geometry that was developed primarily using data from samples of one to five older adults and cadaveric specimens (i.e., Gait2392, 2396Hip, Full Body Running, TLEM2) (Carbone et al., 2015; Delp et al., 1990b; Hamner et al., 2010; Shelburne et al., 2010; Thelen et al., 2012; Yamaguchi and Zajac, 1989). In comparison, only 11 studies used one of the four generic models (i.e., Full Body, LaiArnold2017, 2398Hip, Full Body Squat) developed based on musculotendon parameters derived from MRIs of 24 young, healthy individuals (age range = 12-51) and 21 cadaveric specimens (Handsfield et al., 2014; Rajagopal et al., 2016). Furthermore, the 2396Hip and 2398Hip models, which include additional muscles and updated muscle parameters to be more suitable for hip research, were only used in seven studies.

### 3.3. Model validation

Only eleven (27%) studies included in this review performed model validations (Table 5). The validations were primarily qualitative, and no studies reported any quantitative validation metrics or statistics to quantify the agreement between modeled and experimental HCFs or hip muscle activations (Table 5). Validations for models of more than one patient with a structural hip disorder occurred only within studies examining patient-specific models of people with hip dysplasia and osteoarthritis. Thus, no studies validated generic models for structural hip disorder patients in a sample size greater than one. Furthermore, only seven studies reported that kinematic tracking errors, residual forces, and residual moments were within the recommended limits to verify model quality (Table 5).

**Table 5.** Summary of model validations performed in the included studies.

| Reference | Model | Results Summary |
| --- | --- | --- |
| Buehler et al., 2021 | *Baseline:* Gait2392<br>*Sample:* 16 controls<br>*Patient-specific:* no | *Quantitative:* NR<br>*Qualitative:* “The shape and maximum values of the Orthoload waveforms were similar to the waveforms obtained from our participants, with exception of the [HCF] of the movement leg for the hip abduction exercise, which showed higher values in our participants.” “[HCF] from all exercises in this study showed a reasonable agreement with the values from OrthoLoad.” |
| Lenaerts et al., 2008 | *Baseline:* Gait2392<br>*Iteration*<br>*Sample:* 1 OA patient<br>*Patient-specific:* no | *Quantitative:* NR<br>*Qualitative:* Figures comparing EMG and modeled muscle activity across gait cycle |
| Lenaerts et al., 2009a | *Baseline:* Gait2392<br>*Iteration*<br>*Sample:* 10 OA patients<br>*Patient-specific:* yes | *Quantitative:* Compared to OrthoLoad *in vivo* HCFs, the modeled HCFs had higher magnitudes (3.43 vs. 2.38 BW) and frontal plane inclination angles (33° vs. 13°)<br>*Qualitative:* “The presence of pathological hip kinematics ... contributes to the observed differences [in HCFs]” |
| Gaffney et al., 2020 | *Baseline:* LaiArnold2017<br>*Sample:* 2 HD patients<br>*Patient-specific:* yes | *Quantitative:* RF <10 N for most of gait cycle, max anterior and superior RF <25 N, max medial RF >25 N, RM <50 Nm<br>*Qualitative:* Agreement was shown between modeled and experimental muscle activations, and with the HCFs of the previous modeling study |
| Gaffney et al., 2021 | *Baseline:* Full Body Squat<br>*Sample:* 4 HD patients<br>*Patient-specific:* yes | *Quantitative:* RF: anterior = 21.3 (10.2) N; superior = 23.5 (8.3) N; lateral = 29.5 (19.2) N<br>*Qualitative:* “Model estimated activation timing qualitatively agreed with experimentally collected [EMG]” |
| Harris et al., 2017 | *Baseline:* 2396Hip<br>*Sample:* 1 control<br>*Patient-specific:* yes | *Quantitative:* RF <0.05 BW, RM < 0.3 Nm/kg<br>*Qualitative:* “Model-based activations qualitatively agreed with EMG signals”<br>“... hip JRFs for controls fell near the upper range of those measured in-vivo with telemeterized implants ... and previous models ...”<br>Kinematics, joint moments, and muscle forces were comparable to previous studies |
| Modenese et al., 2021 | *Baseline:* Full Body<br>*Sample:* 1 control<br>*Patient-specific:* yes | *Quantitative:* Reported peak errors, root mean squared errors, and coefficients of determination for comparisons between modeled and experimentally measured knee contact forces (most accurate models were 5° and 12° of femoral anteversion)<br>*Qualitative:* NR |
| Samaan et al., 2019 | *Baseline:* Gait2392<br>*Iteration*<br>*Sample:* 1 (FAIS patient or control)<br>*Patient-specific:* no | *Quantitative:* Root mean squared errors: pelvic translations <1.1 cm; pelvic rotations <0.60°; angles <1.45°; RF <0.66 %BW; RM <0.52 %BW*Height<br>*Qualitative:* “A good qualitative match was found between the EMG and [computed muscle control] estimated muscle activations” |
| Shepherd et al., 2022 | *Baseline:* LaiArnold2017<br>*Sample:* 14 HD patients<br>*Patient-specific:* yes | *Quantitative:* NR<br>*Qualitative:* “... <u>residual forces</u> and moments were minimized to recommended levels...”<br>“Muscle activation timing during gait was checked for qualitative agreement with surface EMG signals...” |
| Shepherd et al., 2023 | *Baseline:* 2396Hip<br>*Sample:* NR<br>*Patient-specific:* yes | *Quantitative:* NR<br>*Qualitative:* “...minimizing residual forces and moments as well as comparing surface electromyography data to predicted muscle activations based on ON/OFF timing and root mean square errors ...” |
| Song et al., 2020 | *Baseline:* 2398Hip<br>*Sample:* 15 HD patients, 15 controls<br>*Patient-specific:* yes | *Quantitative:* Tracking errors = <2 cm; RF <0.025 BW; RM <0.4 Nm/kg<br>*Qualitative:* “Model-estimated muscle activation qualitatively agreed with EMG timings.” “Hip muscle forces and JRFs were in ranges similar to recent subject-specific modeling studies” |

NR = not reported; OA = osteoarthritis; HD = hip dysplasia; FAIS = femoroacetabular impingement syndrome; HCF = hip contact force; EMG = electromyography; BW = body weight; RF = residual forces; RM = residual moments; JRF = joint reaction force.

Figure 2 provides an overview of the original and subsequently developed generic OpenSim models and validations, which we were able to retrieve from the references in the included studies and the software’s documentation. These models were also validated using primarily qualitative methods and with samples of one to ten healthy controls (Figure 2). The most common experimental data used was muscle activations from young, healthy adults and cadaveric muscle moment arm data from older adults (Catelli et al., 2019b; Delp et al., 1990b; Hamner et al., 2010; Lai et al., 2017; Liu et al., 2008; Rajagopal et al., 2016; Shelburne et al., 2010).

**Fig. 2.**
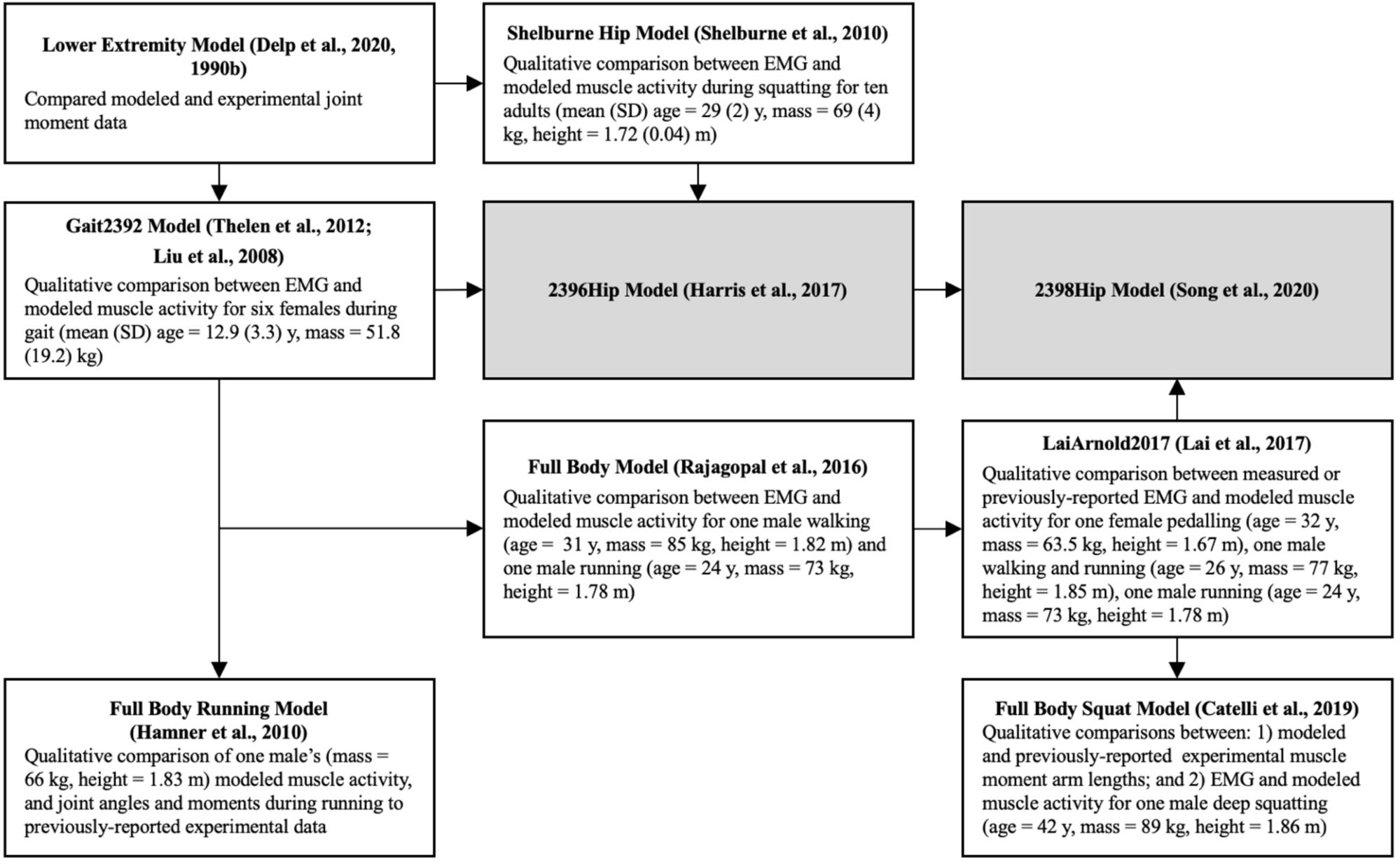
Overview of sequentially developed generic OpenSim models commonly used in the included studies of structural hip disorders showing validations of the original and subsequent model iterations. EMG = electromyography; y = years. Grey boxes indicate models described in studies included in this review (Table 5).

### 3.4. Methods studies

Sixteen studies compared different modeling and simulation methods in the context of structural hip disorders (Table 3). Four of these studies also included analysis of the effect of hip conditions (e.g., patients versus healthy controls) (Table 1). Eleven studies compared the level of model rigid body geometry personalization. In general, there were three levels of model personalization: i) generic models (i.e., geometry scaled based on measures of surface marker locations); ii) generic models deformed to match with imaging measures of patients’ femoral version angle, neck-shaft angle, and/or neck length; and iii) generic models with rigid bodies replaced by fully patient-specific three-dimensional (3D) geometries derived from magnetic resonance imaging (MRI) or computed tomography (CT) scans (Table 3). A subset of these model geometry personalization studies investigated statistical shape modeling (n = 1) and hip joint center (HJC) estimation methods (n = 1). Four studies also performed sensitivity analyses to determine the effect of modeling different femoral version angles (n = 3), femoral neck-shaft angles (n = 2), muscle weakness (n = 2), and femoral neck length (n = 1) (Table 3). Overall, studies showed that incorporating fully patient-specific 3D pelvis and femur geometries and image-based HJCs resulted in different muscle moment arms, muscle forces, and HCFs than those quantified using generic models (Table 3). There were conflicting results regarding whether deforming generic model geometry to match individuals’ measurements (e.g., angle) is an appropriate method to improve model accuracy while minimizing computational cost (Table 3).

Different control models and the use of muscle strength scaling were also investigated. One study demonstrated that EMG-informed modeling improved the estimation of muscle co-contraction in osteoarthritis patients compared to static optimization (Table 3). In addition, one study examined the effect of two methods to scale the model’s maximum isometric muscle forces, which showed that there was either no effect on the HCFs or the model lacked sufficient strength required to generate the joint moments observed during the dynamic tasks (Table 3).

### 3.5. Application studies

Thirty-one studies applied a single modeling workflow to answer a research question related to a structural hip disorder (Table 4). Fourteen studies used the models to compare outputs between hip conditions, four studies compared pre- and post-treatment outputs in patients (e.g., surgical treatment or cued gluteal muscle activation), and five evaluated both a hip condition and a treatment (Table 1). In addition, four studies modified the models to simulate surgical treatment or rehabilitation. Three studies collected data from healthy controls only to investigate a variable related to a structural hip disorder (e.g., HCFs during hip-focused rehabilitation exercises or the effect of muscle weakness on HCFs). Lastly, one study evaluated osteoarthritis patients only to determine the HCFs in response to different rehabilitation exercises.

The rigid body geometry of generic models was most commonly modified to represent an individual using surface marker-based scaling with (n = 14) or without (n = 10) additional personalization from medical imaging. In addition, one study used statistical shape modeling. However, four studies did not report how the models’ rigid bodies were scaled or personalized to represent individual participants. Of the studies that only used surface marker-based scaling, two reported adjusting the HJCs using regression equations. Of the studies that incorporated medical imaging to create patient-specific models, nine implemented image-derived pelvis 3D geometries, eight of which also included femurs. In addition, patient-specific HJCs were typically determined by a least-squares approach to establish the centre of a sphere fit to either the acetabulum or the head of the femur. The remaining studies used imaging data to verify weightings of pelvis markers (n = 3), create pelvis scale factors (n = 1), or deform the generic femur geometry to match the patient-specific femoral version angle (n = 1).

Musculotendon units were also personalized in thirteen application studies (Table 4). For studies that did not replace the generic model geometry, the maximum isometric muscle forces were scaled to the participants’ mass and/or height (n = 3), and the muscle fiber and tendon slack lengths were optimized (n = 2). When image-derived 3D geometries were used to create patient-specific models, the MRIs and knowledge of bony landmarks indicating muscle origin and insertion locations were used to update the muscle paths and attachments. Only one study that used patient-specific models scaled the baseline generic models’ maximum isometric muscle forces to experimental torque data.

### 3.6. Experimental data

Most of the studies collected experimental data which served as model inputs to simulate movement. Motion capture data was collected with various optical tracking systems (n = 46), and ground reaction forces were measured with either force plates (n = 37) or instrumented treadmills (n = 6), (Table 6). Table 6 shows the wide variability in kinematic and kinetic data collection methods across studies. In addition, there was a notable lack of reporting of data collection and signal processing techniques (Table 6). For example, seven studies reported using a 70-marker set (Table 6); however, we could not retrieve information about marker locations from any of these studies or references. Multiple studies did not report the sampling rate for kinematic (n = 11) and kinetic data (n = 10) or how they filtered kinematic (n = 22) and kinetic (n = 19) data. The 11 studies that collected EMG data also demonstrated variability in the signal processing techniques (Table 7). Furthermore, more than a third of EMG studies did not include information on how the EMG signals were filtered (n = 3) or normalized (n = 2).

**Table 6.** Kinematic and kinetic instrumentation and signal processing information for the included studies.

| Reference | Kinematics |  |  |  |  | Kinetics |  |  |  |
| --- | --- | --- | --- | --- | --- | --- | --- | --- | --- |
|  | Cameras | Marker Set | SR (Hz) | Filter | FC (Hz) | Force Plates | SR (Hz) | Filter | FC (Hz) |
| Alexander et al., 2022 | NR | Plug-in gait | 200 | 2<sup>nd</sup> order LPB | 5 | NR | 1000 | 2<sup>nd</sup> order LPB | 12 |
| Bahl et al., 2019 | 10 | 12 markers + 4 rigid clusters | 100 | NR | NR | - | - | - | - |
| Bahl et al., 2020 | 10 | 6DoF | 100 | 2<sup>nd</sup> order LPB | 6 | 2 | 2000 | 2<sup>nd</sup> order LPB | 10 |
| Bosmans et al., 2014 | 8 | SIMM | 100 | NR | NR | 2 | 1500 | NR | NR |
| Bosmans et al., 2016 | 8 | SIMM | 100 | NR | NR | 2 | 1500 | NR | NR |
| Buehler et al., 2021 | 12 | 21 markers + 5 rigid clusters | 100 | NR | NR | 2 | 1000 | NR | NR |
| Cannon et al., 2023 | 11 | 24 markers + 6 rigid clusters | 250 | 2<sup>nd</sup> order LPB | 6 | 2 | 1500 | 2<sup>nd</sup> order LPB | 12 |
| Carriero et al., 2014 | 7 | Helen-Hayes | 50 | NR | NR | 2 | NR | NR | NR |
| Catelli et al., 2019 | 10 | UOMAM | 200 | 4<sup>th</sup> order LPB | 6 | 2 | 1000 | 4<sup>th</sup> order LPB | 6 |
| Catelli et al., 2020 | 10 | UOMAM | 200 | 4<sup>th</sup> order LPB | 6 | 2 | 1000 | 4<sup>th</sup> order LPB | 6 |
| Catelli et al., 2021 | 10 | Modified Plug-in gait | 200 | 4<sup>th</sup> order LPB | 6 | 3 | 1000 | 4<sup>th</sup> order LPB | 6 |
| Choi et al., 2011 | 7 | NR | NR | NR | NR | 2 | NR | NR | NR |
| Diamond et al., 2020 | 12 | 51 markers | 200 | 2<sup>nd</sup> order LPB | 6 | 2 | 1000 | 2<sup>nd</sup> order LPB | 6 |
| De Pieri et al., 2021 | 13 | Institute for Biomechanics | 200 | 4<sup>th</sup> order LPB | 10 | 3 | 1000 | 4<sup>th</sup> order LPB | 20 |
| Foucher et al., 2008 | 4 | 6 markers | NR | NR | NR | 1 | NR | NR | NR |
| Fuller and Winters, 1993 | NR | 14 markers | NR | NR | NR | 2 | NR | NR | NR |
| Gaffney et al., 2020 | NR | 70 markers | 100 | 4<sup>th</sup> order LPB | 8 | Treadmill | 2000 | 4<sup>th</sup> order LPB | 30 |
| Gaffney et al., 2021 | 10 | 70 markers | 100 | 4<sup>th</sup> order LPB | 8 | NR | 2000 | 4<sup>th</sup> order LPB | 20 |
| Harris et al., 2017 | 10 | Modified Helen-Hayes | 100 | LPB | 6 | 4 | 1000 | LPB | 20 |
| Hoang et al., 2019 | 12 | 51 markers | 200 | 2<sup>nd</sup> order LPB | 6 | 2 | 1000 | 2<sup>nd</sup> order LPB | 6 |
| Lenaerts et al., 2008 | NR | NR | NR | NR | NR | NR | NR | NR | NR |
| Lenaerts et al., 2009a | 8 | Plug-in gait | NR | NR | NR | 2 | NR | NR | NR |
| Lenaerts et al., 2009b | 6 | Plug-in gait | NR | NR | NR | 2 | NR | NR | NR |
| Meyer et al., 2018 | 15 | Plug-in gait | 100 | WF* | - | 2 | 1500 | 2<sup>nd</sup> order LPB | 6 |
| Modenese et al., 2021 | NR | NR | NR | NR | NR | NR | NR | NR | NR |
| Ng et al., 2017 | 10 | UOMAM | NR | WF* | - | 2 | NR | 4<sup>th</sup> order LPB | 6 |
| Ng et al., 2018 | 10 | UOMAM | 200 | 4<sup>th</sup> order LPB | 6 | 2 | 1000 | 4<sup>th</sup> order LPB | 6 |
| Passmore et al., 2018 | 10 | Plug-in gait | 100 | WF* | - | NR | 1000 | WF* | - |
| Samaan et al., 2019 | 10 | 29 markers + 4 rigid clusters | 250 | 4<sup>th</sup> order LPB | 6 | 2 | 1000 | 4<sup>th</sup> order LPB | 50 |
| Savage et al., 2022 | 12 | GUFBMS | 120 or 100 | 2<sup>nd</sup> order LPB | 6 | 2 | 1200 or 1000 | 2<sup>nd</sup> order LPB | 6 |
| Scheys et al., 2008 | 8 | SIMM | NR | NR | NR | - | - | - | - |
| Scheys et al., 2011 | 8 | SIMM | NR | NR | NR | - | - | - | - |
| Shepherd et al., 2022 | 10 | 70 markers | 100 | 4<sup>th</sup> order LPB | 8 | Treadmill | 2000 | 4<sup>th</sup> order LPB | 6 |
| Shepherd et al., 2023 | 10 | 70 markers | 100 | NR | NR | Treadmill | 2000 | NR | NR |
| Song et al., 2019 | NR | Modified Helen-Hayes | 100 | 4<sup>th</sup> order LPB | 6 | NR | 1000 | 4<sup>th</sup> order LPB | 20 |
| Song et al., 2020 | 10 | 70 markers | 100 | 4<sup>th</sup> order LPB | 8 | Treadmill | 2000 | 4<sup>th</sup> order LPB | 6 |
| Song et al., 2021 | 10 | 70 markers | 100 | LPB | 8 | Treadmill | 2000 | LPB | 6 |
| Song et al., 2022 | 10 | 70 markers | 100 | LPB | 8 | 2 | 2000 | LPB | 10 |
| Tateuchi et al., 2021 | 8 | Plug-in gait | 100 | 4<sup>th</sup> order LPB | 6 | NR | 1000 | 4<sup>th</sup> order LPB | 6 |
| Vandekerckhove et al., 2021 | 12 | Plug-in gait | 100 | NR | NR | 2 | 1000 | NR | NR |
| Van Rossom et al., 2023 | 13 | Modified Plug-in gait | 100 | NR | NR | NR | 1000 | NR | NR |
| Wesseling et al., 2016 | NR | Plug-in gait | 100 | NR | NR | 2 | 1500 | NR | NR |
| Wesseling et al., 2018 | NR | Plug-in gait | 100 | NR | NR | 2 | 1500 | NR | NR |
| Wesseling et al., 2019a | 8 | SIMM | 100 | NR | NR | 2 | 1500 | NR | NR |
| Wesseling et al., 2019b | 10 | Plug-in gait | 100 | NR | NR | 2 | 1000 | NR | NR |
| Wu et al., 2023 | NR | NR | NR | NR | NR | Treadmill | NR | NR | NR |

NR = not reported; SR = sampling rate; FC = filter cut-off; LPB = low-pass Butterworth; WF = Woltring filter. \*Mean squared error = 15mm<sup>2</sup>

**Table 7.**
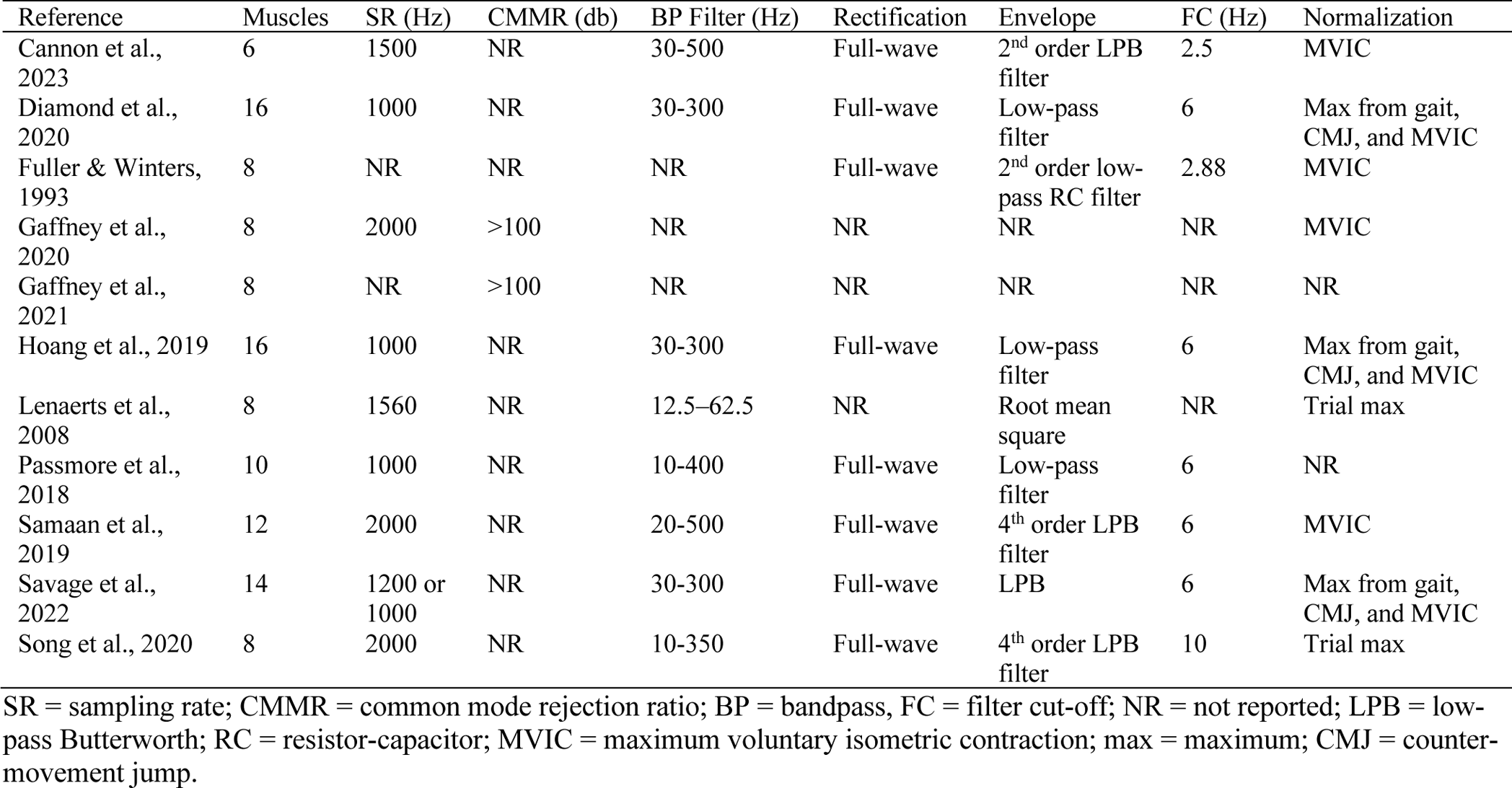
Electromyography instrumentation and signal processing information for the included studies.

### 3.7. Simulation workflow

The majority of the studies used a standard MSK modeling workflow starting by first calculating joint angles from motion capture data using inverse kinematics. Then, an inverse dynamics tool was used to calculate external joint moments and forces based on ground reaction forces and kinematics. Next, individual muscle forces were estimated, followed by joint contact forces based on the external joint forces and the individual muscle forces. The most common control model used to estimate muscle forces for the methods studies was static optimization (n = 10), while one study used computed muscle control (Table 3), excluding the study that investigated different control models. For the application studies, 21 studies used static optimization, three used EMG-assisted models, and three used computed muscle control models (Table 4).

## 4. Discussion

Forty-seven studies were identified in this scoping review that used MSK models as a tool to research structural hip disorders. The hip disorders investigated in these studies included osteoarthritis, hip dysplasia, cerebral palsy-associated femoral deformities, FAIS, and idiopathic femoral version deformities. The models were primarily used to quantify HCFs and muscle forces or activations in response to dynamic tasks *in vivo.* Generic OpenSim models were most commonly used. However, there were a lack of quantitative methods used to validate these generic models, especially in the context of structural hip disorders. In addition, the use of recommended methods from studies that compared various modeling and simulation techniques was limited in studies that used the models to answer clinical research questions. Lastly, the wide variability and under-reporting of data collection, data processing, and modeling methods present barriers to comparing studies and limit study reproducibility.

### 4.1. Model development and validation

Retrieving information regarding model development and validation results for the generic models presented a challenge due to missing detailed descriptions of models and model iteration information that was common in structural hip disorder modeling studies. Based on our knowledge, the common generic models reported in the included studies were primarily developed based on data from older adults and cadaveric specimens, then most commonly validated using experimental muscle activation data from young, healthy adults. In contrast, the populations in the included studies demonstrated large heterogeneity in demographics and activity levels. The validations performed in the included studies of structural hip disorders were also primarily limited to qualitative assessments of experimental and model data agreement. While validation guidelines have been shared in terms of the timing of muscle activations and agreement with experimental HCFs (Hicks et al., 2015), these quantitative validation methods have not been widely adopted in the modeling of structural hip disorders. Therefore, additional validation is required to assess the representativeness of the generic models to quantify hip joint mechanics in the specific populations of interest. Ongoing validation of the models shared by researchers would generate important, up-to-date knowledge of the accuracy of current models.

Updated validation thresholds are needed that are specific to populations with structural hip disorders. For example, it is currently recommended that modeled HCFs fall within two standard deviations of experimental HCFs (Hicks et al., 2015). However, experimental HCFs are only available from instrumented hip prostheses in total hip arthroplasty (THA) patients (Bergmann et al., 2016, 2001). Therefore, it is likely that previously-reported differences between the model and experimental HCFs are more attributable to the inherent demographic and physical differences between older THA patients and other populations (e.g., young active adults with FAIS) rather than the errors within the models themselves. Specifically, when young, healthy adults squat to maximum depth, their HCFs have been noted to reach values greater than two standard deviations higher than the older THA patients; however, when looking at the HCFs of the young adults when at the mean knee flexion angle of the THA patients (i.e., 71°), the HCFs are lower than the THA patients and within two standard deviations (Harrington and Burkhart, 2023). These findings suggest a need for continued discussions to determine the level of modeling validation required to answer clinically relevant research questions. It is also evident that more representative experimental data sets are required.

### 4.2. Modeling and simulation methods

Overall, the studies included in this review that compared generic and patient-specific modeling methods recommended using patient-specific models. This recommendation was based on their results demonstrating differences in HCFs and muscle moment arm lengths between models with different levels of geometry personalization. However, due to the challenges of measuring *in vivo* HCFs for validation and a lack of validation studies quantitatively comparing experimental and modeled muscle activations and parameters, it is unclear whether the patient-specific models are truly more accurate. This likely contributes to the lack of adoption of patient-specific modeling in application studies, with only 36% of studies using image-derived 3D geometries.

A potential concern with patient-specific modeling methods is that they rely on semi-manual adjustment of muscle paths to match imaging data and anatomical descriptions of muscle attachments from the literature (Gaffney et al., 2021; Scheys et al., 2011). Thus, it is suggested that there is a potential for inter-operator and inter-patient errors when creating patient-specific models (Benemerito et al., 2022; Killen et al., 2021); however, the amount of error in modeling of different structural hip disorders is unclear and not reported. In addition, since MRIs only capture static positions, there is uncertainty regarding the accuracy of MRI-based muscle paths through the entire joint range of motion (Song et al., 2020). Muscle moment arms and lines of action can be a significant source of error in model muscle activation and HCF estimations and may primarily account for differences in output between patient-specific and generic MSK models (Wesseling et al., 2019a). Given the cost of acquiring MRIs or CT scans for study participants and the time required to generate patient-specific models, it is important to determine the level of model geometry personalization and accuracy required to answer given research questions. Quantitative validations are also essential to help establish this threshold for accuracy.

The methods study conducted by Hoang et al. (2019) established that EMG-informed models better predict muscle co-activation compared to static optimization; however, only one subsequent study has used this control model (Diamond et al., 2020). Considering increased muscle co-activation is expected in those with hip osteoarthritis, the under-estimation of muscle co-activation is important in modeling research that aims to quantify muscle forces within this population. EMG-informed models also consistently estimated significantly higher HCFs than static optimization across the gait cycle (Hoang et al., 2019). Given that most studies applying models to study a specific hip disorder are interested in the relative difference in HCFs between populations or time points, knowing the error between control modeling methods can help to evaluate whether these modeling decisions would impact the overall interpretation of study results. Future comparisons between HCFs estimated with static optimization versus EMG-informed models in populations with other hip disorders with or without abnormal patterns of muscle co-contraction will help to support researchers’ decisions about control models.

The current review also highlighted that MSK modeling of structural hip disorders is still predominately used to investigate tasks with limited hip range of motion (e.g., gait). Studies investigating young adult hip disorders, such as FAIS and hip dysplasia, were a notable proportion of the articles included in this review. Given that these populations are generally young and active, tasks such as gait may not be sufficient to comprehensively evaluate abnormal hip mechanics using MSK modeling. This divide may be present because modeling of the hip has primarily been focused on individuals with cerebral palsy or hip osteoarthritis, who typically present with more severe gait deficits than those shown in FAIS or hip dysplasia patients (Bosmans et al., 2014; Catelli et al., 2019a; Diamond et al., 2020; Harris et al., 2017; Meyer et al., 2018). Models that simulate deep squats (Catelli et al., 2019b; Lai et al., 2017; Song et al., 2020) have been recently developed but have yet to be widely adopted. In addition, the 2396Hip model was recently modified and quantitatively validated in healthy controls to simulate dynamic tasks with increased multiplanar hip joint ranges of motions (Harrington and Burkhart, 2023). Further development and validation of models to investigate a wider variety of clinically relevant tasks is an important area of opportunity and advancement for MSK modeling of structural hip disorders.

### 4.3. Study reproducibility

The results of this scoping review highlight inconsistencies in the reporting of modeling and simulation method details. For example, it was common for studies using an iteration of the Gait2392 model to cite the original article describing the original Lower Extremity Model developed by Delp et al. (1990b) without specifying which iteration of the model they were utilising. This is challenging because the same citation was used in studies that had implemented models with different degrees of freedom or numbers of musculotendon actuators. There was also minimal reporting of the details at other stages of the modeling and simulation workflow. For example, few studies clarified how many scale factors were used to scale the models and which markers were used to calculate the scale factors. In addition, only seven studies reported the residual errors for marker location for scaling or tracking during inverse kinematics and the residual forces or moments during inverse dynamics. More detailed reporting guidelines should be established and enforced to help researchers assess their modeling studies and improve reproducibility.

Reporting the instrumentation specifications and data collection and processing techniques was inconsistent across studies. For example, many studies did not report sufficient details of the marker set used, the number of cameras, or the sampling rate at which data was collected. Previous studies have shown that these data collection parameters can strongly influence the results of biomechanical analyses. Specifically in modeling, Mantovani and Lamontagne (2017) demonstrated significant differences in joint angles calculated in OpenSim between three marker set configurations. Two of these configurations were commonly used in the studies included in this review (Plug-in-Gait and the University of Ottawa Motion Analysis Model). However, it remains unclear how different marker sets may influence other model outputs such as the commonly estimated HCFs. Studies have also shown variations in biomechanical outcomes between labs, specifically with joint moments when a standardized protocol is not being used (Benedetti et al., 2013; Kaufman et al., 2016). Furthermore, the largest inter-laboratory inconsistency in gait measurements was pelvis anthropometric measures, which are critical to the most common marker-based scaling and HJC estimation methods used in the studies included in this review. Enhancing the clarity in the reporting of data collection and processing techniques will likely make it easier to assess the accuracy and reproducibility of the results.

It was unclear how the raw experimental data was processed in 48% and 44% of studies using kinematic and kinetic data, respectively. In addition, most of the studies that reported how the data was filtered did not describe the rationale behind choosing a specific filter cut-off frequency. Cut-off frequency choice is an important signal processing consideration, particularly when filtering data that will be input into MSK models. For example, Tomescu et al. (2018) compared different marker and ground reaction force filter cut-off frequency conditions and quantified significant differences in model-predicted muscle forces, joint contact forces, and residual forces and moments. A greater emphasis should be placed on reporting the signal processing details of input data for models as it can profoundly impact the results of modeling studies. While there are unavoidable uncertainties and assumptions required in MSK modeling, good and transparent data collection and processing methods can be easily implemented to help improve researchers’ and clinicians’ ability to interpret the clinical relevance of modeling simulation results.

## 5. Conclusion

MSK models are a powerful tool to provide insight into factors that are typically not feasible to measure *in vivo,* such as HCFs and deep muscle activations. Furthermore, they represent an opportunity to simulate the effects of treatments and generate inputs for different scales of models, such as finite element models, to address broader research questions. This scoping review identified that there is limited quantitative data available to fully understand the accuracy of MSK models, specifically in the context of structural hip disorders. More stringent validation studies will also help researchers reach a consensus on the level of model personalization and the optimal control models required for MSK models that are focused on structural hip disorders. Increased transparency in reporting data collection, signal processing, and modeling methods is needed to increase study reproducibility and allow researchers and clinicians to better assess modeling study results.

## Data Availability

All data produced in the present work are contained in the manuscript.

## CRediT Authorship Contribution Statement

Margaret S. Harrington: Conceptualization, Methodology, Formal analysis, Investigation, Data curation, Writing – original draft, Writing –review & editing, Supervision, Project administration. Stefania D. F. Di Leo: Formal analysis, Investigation, Data Curation, Writing – review & editing. Courtney A. Hlady: Formal analysis, Investigation, Writing – review & editing. Timothy A. Burkhart: Methodology, Resources, Writing – review & editing, Supervision, Project administration, Funding acquisition.

## Acknowledgments

The authors would like to thank Julia Martyniuk for their valuable support in developing the search strategy. The authors would also like to thank Pratham Singh and Lucy Tempest for their assistance screening the titles, abstracts, and full-text reports for inclusion and exclusion. This work was supported by the Natural Sciences and Engineering Research Council of Canada

## Appendix A

Databases were searched twice:

- First search: October 20^th^, 2021
- Second search: July 3^rd^, 2023

**Table A1.**
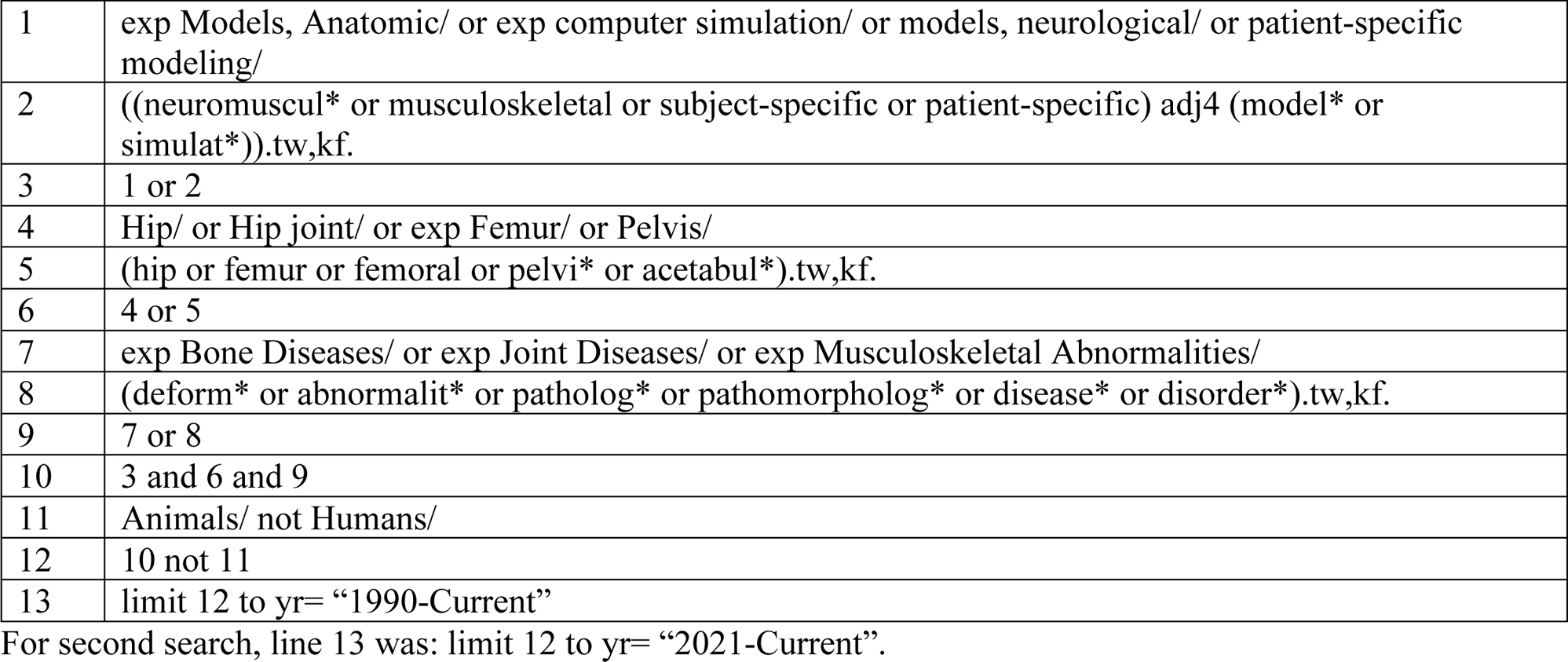
Medline Search Strategy.

|  |  |
| --- | --- |
| 1 | exp Models, Anatomic/ or exp computer simulation/ or models, neurological/ or patient-specific modeling/ |
| 2 | ((neuromuscul* or musculoskeletal or subject-specific or patient-specific) adj4 (model* or simulat*)).tw,kf. |
| 3 | 1 or 2 |
| 4 | Hip/ or Hip joint/ or exp Femur/ or Pelvis/ |
| 5 | (hip or femur or femoral or pelvi* or acetabul*).tw,kf. |
| 6 | 4 or 5 |
| 7 | exp Bone Diseases/ or exp Joint Diseases/ or exp Musculoskeletal Abnormalities/ |
| 8 | (deform* or abnormalit* or patholog* or pathomorpholog* or disease* or disorder*).tw,kf. |
| 9 | 7 or 8 |
| 10 | 3 and 6 and 9 |
| 11 | Animals/ not Humans/ |
| 12 | 10 not 11 |
| 13 | limit 12 to yr= "1990-Current" |

For second search, line 13 was: limit 12 to yr= "2021-Current".

**Table A2.** Web of Science Search Strategy.

|  |  |
| --- | --- |
| 1 | TS=((neuromuscul* or musculoskeletal or subject-specific or patient-specific) near/3 (model* or simulat*)) |
| 2 | TS=(hip or femur or femoral or pelvi* or acetabul*) |
| 3 | TS=(deform* or abnormalit* or patholog* or pathomorpholog* or disease* or disorder*) |
| 4 | 1 and 2 and 3 |
| 5 | TS=(Animal* not Human*) |
| 6 | 4 not 5 |
| 7 | Publication date = Jan 1, 1990 - Oct 20, 2021 |

For second search, line 7 was: Publication date = Oct 20, 2021 – July 3, 2021

## Appendix B

Modified quality assessment checklist originally developed by Moissenet et al. (2017):

Q1: Are the research objectives clearly stated?
Q2: Is the study design clearly described?
Q3: Is the scientific context clearly explained?
Q4: Is the musculoskeletal model adequately described?
Q5: Were the model alterations clearly described?
Q6: Is the model for joint contact force estimation adequately described?
Q7: Were participant characteristics adequately described?
Q8: Were movement tasks, equipment design, and setup clearly defined?
Q9: Were relevant instrumentation specifications and signal processing techniques described?
Q10: Were the statistical methods justified and appropriately described (other than descriptive statistics)?
Q11: Were the direct results easily interpretable?
Q12: Were the main outcomes clearly stated and supported by the results?
Q13: Were the limitations of the study clearly described?
Q14: Were key findings supported by other literature?
Q15: Were conclusions drawn from the study clearly stated?

**Table B1.** Quality assessment of included studies using a modified checklist developed by Moissenet et al. (2017) for biomechanical research.

| Reference | Q1 | Q2 | Q3 | Q4 | Q5 | Q6 | Q7 | Q8 | Q9 | Q10 | Q11 | Q12 | Q13 | Q14 | Q15 | Score (%) |
| --- | --- | --- | --- | --- | --- | --- | --- | --- | --- | --- | --- | --- | --- | --- | --- | --- |
| Alexander et al., 2022 | 2 | 2 | 2 | 2 | 2 | 2 | 1 | 1 | 2 | 1 | 1 | 2 | 2 | 1 | 2 | 83 |
| Bahl et al., 2019 | 2 | 2 | 2 | 2 | 1 | - | 2 | 2 | 1 | 2 | 1 | 1 | 2 | 2 | 2 | 86 |
| Bahl et al., 2020 | 2 | 2 | 2 | 1 | 2 | 1 | 2 | 1 | 1 | 2 | 1 | 2 | 2 | 2 | 2 | 83 |
| Bosmans et al., 2014 | 1 | 2 | 2 | 1 | 2 | 1 | 2 | 2 | 1 | 2 | 1 | 2 | 2 | 2 | 2 | 83 |
| Bosmans et al., 2016 | 1 | 2 | 2 | 1 | 2 | - | 2 | 1 | 0 | 2 | 1 | 2 | 2 | 2 | 2 | 79 |
| Buehler et al., 2021 | 2 | 2 | 2 | 1 | 1 | 0 | 1 | 2 | 1 | 2 | 1 | 2 | 2 | 1 | 2 | 73 |
| Cannon et al., 2023 | 2 | 2 | 2 | 1 | 1 | 1 | 2 | 2 | 2 | 1 | 2 | 2 | 2 | 2 | 2 | 87 |
| Carriero et al., 2014 | 2 | 2 | 2 | 1 | - | 1 | 1 | 2 | 1 | - | 1 | 2 | 2 | 1 | 2 | 77 |
| Catelli et al., 2019a | 2 | 2 | 2 | 2 | 2 | 1 | 2 | 2 | 1 | 2 | 2 | 1 | 2 | 2 | 2 | 90 |
| Catelli et al., 2020 | 2 | 2 | 2 | 2 | 2 | 1 | 2 | 2 | 1 | 2 | 2 | 2 | 2 | 2 | 2 | 93 |
| Catelli et al., 2021 | 2 | 2 | 2 | 2 | 2 | 1 | 1 | 2 | 1 | 2 | 2 | 1 | 2 | 2 | 1 | 83 |
| Choi et al., 2011 | 1 | 2 | 2 | 1 | - | - | 2 | 2 | 1 | 1 | 1 | 2 | 1 | 1 | 2 | 73 |
| De Pieri et al., 2021 | 2 | 2 | 2 | 2 | 2 | 2 | 1 | 2 | 1 | 1 | 1 | 2 | 2 | 2 | 1 | 83 |
| Delp et al., 1990 | 2 | 2 | 2 | 1 | 2 | - | 1 | 1 | 1 | 0 | 2 | 2 | 2 | 2 | 2 | 79 |
| Diamond et al., 2020 | 2 | 2 | 2 | 2 | 2 | 1 | 2 | 1 | 1 | 2 | 2 | 2 | 2 | 2 | 2 | 90 |
| Foucher et al., 2008 | 2 | 2 | 2 | 1 | - | 1 | 2 | 2 | 1 | 2 | 2 | 2 | 2 | 1 | 2 | 86 |
| Fuller and Winters, 1993 | 2 | 2 | 2 | 1 | 1 | 1 | 1 | 1 | 1 | 1 | 1 | 1 | 2 | 2 | 2 | 70 |
| Gaffney et al., 2020 | 2 | 2 | 2 | 1 | 1 | 1 | 2 | 1 | 1 | 2 | 1 | 2 | 2 | 2 | 2 | 80 |
| Gaffney et al., 2021 | 2 | 2 | 2 | 2 | 2 | 1 | 2 | 1 | 1 | 2 | 2 | 2 | 2 | 2 | 2 | 90 |
| Harris et al., 2017 | 2 | 2 | 2 | 2 | 1 | 1 | 2 | 2 | 2 | 2 | 2 | 2 | 2 | 2 | 2 | 93 |
| Hoang et al., 2019 | 2 | 2 | 2 | 2 | 2 | 1 | 2 | 1 | 1 | 2 | 2 | 1 | 2 | 2 | 2 | 87 |
| Lenaerts et al., 2008 | 2 | 2 | 2 | 2 | 1 | 1 | 2 | 1 | 1 | 2 | 2 | 2 | 2 | 2 | 2 | 87 |
| Lenaerts et al., 2009a | 2 | 2 | 2 | 1 | 1 | 1 | 2 | 1 | 1 | 2 | 2 | 1 | 1 | 2 | 2 | 77 |
| Lenaerts et al., 2009b | 2 | 2 | 2 | 1 | 2 | 1 | 2 | 1 | 1 | 1 | 2 | 2 | 2 | 2 | 2 | 83 |
| Meyer et al., 2018 | 2 | 2 | 2 | 1 | 1 | 1 | 2 | 2 | 1 | 2 | 2 | 2 | 1 | 2 | 1 | 80 |
| Modenese et al., 2021 | 2 | 2 | 2 | 2 | 0 | 2 | 1 | 1 | 1 | 2 | 2 | 2 | 1 | 2 | 2 | 80 |
| Ng et al., 2017 | 2 | 2 | 2 | 2 | 1 | 2 | 2 | 2 | 1 | 1 | 2 | 2 | 2 | 2 | 2 | 90 |
| Ng et al., 2018 | 2 | 2 | 2 | 2 | 1 | 1 | 2 | 2 | 1 | 2 | 2 | 2 | 2 | 2 | 2 | 90 |
| Passmore et al., 2018 | 2 | 2 | 2 | 2 | 1 | 2 | 2 | 1 | 1 | 2 | 2 | 2 | 2 | 2 | 1 | 87 |
| Samaan et al., 2019 | 2 | 2 | 2 | 2 | 2 | - | 2 | 2 | 1 | 2 | 1 | 1 | 2 | 2 | 2 | 89 |
| Savage et al., 2022 | 1 | 2 | 2 | 2 | 1 | 2 | 2 | 1 | 1 | 2 | 1 | 2 | 2 | 2 | 2 | 83 |
| Scheys et al., 2008 | 1 | 2 | 2 | 1 | 2 | - | 2 | 1 | 1 | 2 | 1 | 2 | 1 | 2 | 2 | 79 |
| Scheys et al., 2011 | 1 | 2 | 2 | 1 | 2 | - | 2 | 1 | 1 | 2 | 1 | 2 | 2 | 2 | 1 | 79 |
| Shepherd et al., 2022 | 2 | 2 | 2 | 2 | 2 | 2 | 2 | 2 | 1 | 2 | 1 | 2 | 2 | 2 | 2 | 93 |
| Shepherd et al., 2023 | 2 | 2 | 2 | 2 | 2 | 2 | 2 | 2 | 1 | 2 | 1 | 2 | 2 | 2 | 2 | 93 |
| Song et al., 2019 | 2 | 2 | 2 | 2 | 2 | 1 | 2 | 2 | 1 | 2 | 2 | 2 | 2 | 2 | 2 | 93 |
| Song et al., 2020 | 2 | 2 | 2 | 2 | 2 | 1 | 2 | 2 | 2 | 2 | 1 | 2 | 2 | 2 | 2 | 93 |
| Song et al., 2021 | 2 | 2 | 2 | 1 | 2 | 1 | 2 | 2 | 2 | 2 | 1 | 2 | 2 | 2 | 2 | 90 |
| Song et al., 2022 | 2 | 2 | 2 | 2 | 1 | 2 | 2 | 1 | 2 | 2 | 2 | 2 | 2 | 2 | 2 | 93 |
| Tateuchi et al., 2021 | 1 | 2 | 2 | 2 | 2 | 1 | 2 | 2 | 1 | 2 | 1 | 2 | 2 | 2 | 2 | 87 |
| Van Rossom et al., 2023 | 1 | 2 | 2 | 1 | 1 | 2 | 1 | 1 | 1 | 1 | 2 | 2 | 2 | 2 | 2 | 77 |
| Vandekerckhove et al., 2021 | 2 | 2 | 2 | 2 | 2 | - | 2 | 1 | 1 | 2 | 2 | 2 | 2 | 2 | 2 | 93 |
| Wesseling et al., 2016 | 1 | 2 | 2 | 2 | 1 | 1 | 2 | 1 | 1 | 0 | 2 | 2 | 2 | 2 | 2 | 77 |
| Wesseling et al., 2018 | 2 | 2 | 2 | 2 | 1 | 1 | 2 | 1 | 1 | 2 | 2 | 1 | 2 | 2 | 2 | 83 |
| Wesseling et al., 2019a | 2 | 2 | 2 | 2 | 2 | 1 | 2 | 1 | 1 | 2 | 2 | 2 | 2 | 1 | 2 | 87 |
| Wesseling et al., 2019b | 2 | 2 | 2 | 1 | 2 | 1 | 2 | 2 | 1 | - | 2 | 2 | 2 | 2 | 2 | 89 |
| Wu et al., 2023 | 1 | 1 | 2 | 1 | 1 | 2 | 2 | 1 | 0 | 1 | 2 | 2 | 2 | 2 | 2 | 73 |

## References

Alexander, N., Brunner, R., Cip, J., Viehweger, E., De Pieri, E., 2022. Increased Femoral Anteversion Does Not Lead to Increased Joint Forces During Gait in a Cohort of Adolescent Patients. Front. Bioeng. Biotechnol. 10. 914990. 10.3389/fbioe.2022.914990

Bahl, J.S., Arnold, J.B., Taylor, M., Solomon, L.B., Thewlis, D., 2020. Lower functioning patients demonstrate atypical hip joint loading before and following total hip arthroplasty for osteoarthritis. J. Orthop. Res. 38 (7), 1550–1558. 10.1002/JOR.24716

Bahl, J.S., Zhang, J., Killen, B.A., Taylor, M., Solomon, L.B., Arnold, J.B., Lloyd, D.G., Besier, T.F., Thewlis, D., 2019. Statistical shape modelling versus linear scaling: Effects on predictions of hip joint centre location and muscle moment arms in people with hip osteoarthritis. J. Biomech. 85, 164–172. 10.1016/j.jbiomech.2019.01.031

Benedetti, M.G., Merlo, A., Leardini, A., 2013. Inter-laboratory consistency of gait analysis measurements. Gait Posture. 38 (4), 934–939. 10.1016/j.gaitpost.2013.04.022

Benemerito, I., Montefiori, E., Marzo, A., Mazzà, C., 2022. Reducing the Complexity of Musculoskeletal Models Using Gaussian Process Emulators. Appl. Sci. 12 (14), 12932. 10.3390/app122412932

Bergmann, G., Bender, A., Dymke, J., Duda, G., Damm, P., 2016. Standardized loads acting in hip implants. PLoS One. 11 (5): e0155612. 10.1371/journal.pone.0155612

Bergmann, G., Deuretzbacher, G., Heller, M., Graichen, F., Rohlmann, A., Strauss, J., Duda, G.N., 2001. Hip contact forces and gait patterns from routine activities. J. Biomech. 34 (7), 859–871. 10.1016/S0021-9290(01)00040-9

Bosmans, L., Jansen, K., Wesseling, M., Molenaers, G., Scheys, L., Jonkers, I., 2016. The role of altered proximal femoral geometry in impaired pelvis stability and hip control during CP gait: A simulation study. Gait Posture. 44, 61–67. 10.1016/j.gaitpost.2015.11.010

Bosmans, L., Wesseling, M., Desloovere, K., Molenaers, G., Scheys, L., Jonkers, I., 2014. Hip Contact Force in Presence of Aberrant Bone Geometry During Normal and Pathological Gait. J. Orthop. Res. 32 (11), 1406–1415. 10.1002/jor.22698

Buehler, C., Koller, W., De Comtes, F., Kainz, H., 2021. Quantifying Muscle Forces and Joint Loading During Hip Exercises Performed With and Without an Elastic Resistance Band. Front. Sports Act. Living. 3, 695383. 10.3389/fspor.2021.695383

Cannon, J., Kulig, K., Weber, A.E., Powers, C.M., 2023. Gluteal activation during squatting reduces acetabular contact pressure in persons with femoroacetabular impingement syndrome: A patient-specific finite element analysis. Clin. Biomech. 101, 105849. 10.1016/J.CLINBIOMECH.2022.105849

Carriero, A., Zavatsky, A., Stebbins, J., Theologis, T., Lenaerts, G., Jonkers, I., Shefelbine, S.J., 2014. Influence of altered gait patterns on the hip joint contact forces. Comput. Methods Biomech. Biomed. Engin. 17 (4), 352–359. 10.1080/10255842.2012.683575

Catelli, D.S., Bedo, B.L.S., Beaulé, P.E., Lamontagne, M., 2021. Pre- and postoperative in silico biomechanics in individuals with cam morphology during stair tasks. Clin. Biomech. 86, 105387. 10.1016/j.clinbiomech.2021.105387

Catelli, D.S., Ng, K.C.G., Kowalski, E., Beaulé, P.E., Lamontagne, M., 2019a. Modified gait patterns due to cam FAI syndrome remain unchanged after surgery. Gait Posture. 72, 135–141. 10.1016/j.gaitpost.2019.06.003

Catelli, D.S., Ng, K.C.G., Wesseling, M., Kowalski, E., Jonkers, I., Beaulé, P.E., Lamontagne, M., 2020. Hip Muscle Forces and Contact Loading During Squatting After Cam-Type FAI Surgery. J. Bone Joint Surg. Am. 102 (Suppl 2), 34–42. 10.2106/JBJS.20.00078

Catelli, D.S., Wesseling, M., Jonkers, I., Lamontagne, M., 2019b. A musculoskeletal model customized for squatting task. Comput. Methods Biomech. Biomed. Engin. 22 (1), 21–24. 10.1080/10255842.2018.1523396

Choi, S.J., Chung, C.Y., Lee, K.M., Kwon, D.G., Lee, S.H., Park, M.S., 2011. Validity of gait parameters for hip flexor contracture in patients with cerebral palsy. J. Neuroeng. Rehabil. 8, 4. 10.1186/1743-0003-8-4

Clohisy, J.C., Keeney, J.A., Schoenecker, P.L., 2005. Preliminary assessment and treatment guidelines for hip disorders in young adults. Clin. Orthop. Relat. Res. 441, 168–179. 10.1097/01.blo.0000193511.91643.2a

Damsgaard, M., Rasmussen, J., Christensen, S.T., Surma, E., de Zee, M., 2006. Analysis of musculoskeletal systems in the AnyBody Modeling System. Simul. Mode.l Pract. Theory. 14 (8), 1100–11. 10.1016/j.simpat.2006.09.001

De Pieri, E., Friesenbichler, B., List, R., Monn, S., Casartelli, N.C., Leunig, M., Ferguson, S.J., 2021. Subject-Specific Modeling of Femoral Torsion Influences the Prediction of Hip Loading During Gait in Asymptomatic Adults. Front. Bioeng. Biotechnol. 9, 679360. 10.3389/fbioe.2021.679360

Delp, S.L., 2020. Lower Extremity Model. SimTK. Retrieved July 3, 2023, from: https://simtk.org/projects/low-ext-model#:~:text=Originally%20developed%20in%20DATE%20by,represent%20the%20human%20lower%20extremity.

Delp, S.L., Bleck, E.E., Zajac, F.E., Bollini, G., 1990a. Biomechanical analysis of the Chiari pelvic osteotomy. Preserving hip abductor strength. Clin. Orthop. Relat. Res. 254, 189–198.

Delp, S.L., Loan, J.P., Hoy, M.G., Zajac, F.E., Topp, E.L., Rosen, J.M., 1990b. An Interactive Graphics-Based Model of the Lower Extremity to Study Orthopaedic Surgical Procedures. IEEE Trans. Biomed. Eng. 37 (8), 757–767. 10.1109/10.102791

Diamond, L.E., Hoang, H.X., Barrett, R.S., Loureiro, A., Constantinou, M., Lloyd, D.G., Pizzolato, C., 2020. Individuals with mild-to-moderate hip osteoarthritis walk with lower hip joint contact forces despite higher levels of muscle co-contraction compared to healthy individuals. Osteoarthritis Cartilage. 28 (7), 924–931. 10.1016/j.joca.2020.04.008

Foucher, K.C., Hurwitz, D.E., Wimmer, M.A., 2008. Do gait adaptations during stair climbing result in changes in implant forces in subjects with total hip replacements compared to normal subjects? Clin. Biomech. 23 (6), 754–761. 10.1016/j.clinbiomech.2008.02.006

Fuller, J.J., Winters, J.M., 1993. Assessment of 3-D Joint Contact Load Predictions During Posturai/Stretching Exercises in Aged Females, Ann. Biomed. Eng. 21 (3), 277–288. 10.1007/BF02368183

Gaffney, B.M.M., Clohisy, J.C., Van Dillen, L.R., Harris, M.D., 2020. The association between periacetabular osteotomy reorientation and hip joint reaction forces in two subgroups of acetabular dysplasia. J. Biomech. 98, 109464. 10.1016/J.JBIOMECH.2019.109464

Gaffney, B.M.M., Harris-Hayes, M., Clohisy, J.C., Harris, M.D., 2021. Effect of simulated rehabilitation on hip joint loading during single limb squat in patients with hip dysplasia. J. Biomech. 116, 110183. 10.1016/j.jbiomech.2020.110183

Hamner, S.R., Seth, A., Delp, S.L., 2010. Muscle contributions to propulsion and support during running. J. Biomech. 43 (14), 2709–2716. 10.1016/j.jbiomech.2010.06.025

Handsfield, G.G., Meyer, C.H., Hart, J.M., Abel, M.F., Blemker, S.S., 2014. Relationships of 35 lower limb muscles to height and body mass quantified using MRI. J. Biomech. 47 (3), 631–8. 10.1016/j.jbiomech.2013.12.002

Harrington, M.S., Burkhart, T.A., 2023. Validation of a musculoskeletal model to investigate hip joint mechanics in response to dynamic multiplanar tasks. J. Biomech. 158:111767. 10.1016/j.jbiomech.2023.111767

Harris, M.D., MacWilliams, B.A., Bo Foreman, K., Peters, C.L., Weiss, J.A., Anderson, A.E., 2017. Higher medially-directed joint reaction forces are a characteristic of dysplastic hips: A comparative study using subject-specific musculoskeletal models. J. Biomech. 54, 80–87. 10.1016/j.jbiomech.2017.01.040

Hicks, J.L., Uchida, T.K., Seth, A., Rajagopal, A., Delp, S.L., 2015. Is My Model Good Enough? Best Practices for Verification and Validation of Musculoskeletal Models and Simulations of Movement. J. Biomech. Eng. 137 (2), 020905. 10.1115/1.4029304

Hoang, H.X., Diamond, L.E., Lloyd, D.G., Pizzolato, C., 2019. A calibrated EMG-informed neuromusculoskeletal model can appropriately account for muscle co-contraction in the estimation of hip joint contact forces in people with hip osteoarthritis. J. Biomech. 83, 134–142. 10.1016/j.jbiomech.2018.11.042

Husen, M., Leland, D.P., Melugin, H.P., Poudel, K., Hevesi, M., Levy, B.A., Krych, A.J., 2023. Progression of Osteoarthritis at Long-term Follow-up in Patients Treated for Symptomatic Femoroacetabular Impingement With Hip Arthroscopy Compared With Nonsurgically Treated Patients. Am. J. Sports Med. 51 (11), 2986–95. 10.1177/03635465231188114

Kaufman, K., Miller, E., Kingsbury, T., Russell Esposito, E., Wolf, E., Wilken, J., Wyatt, M., 2016. Reliability of 3D gait data across multiple laboratories. Gait Posture. 49, 375–381. 10.1016/j.gaitpost.2016.07.075

Kiger, M.E., Varpio, L., 2020. Thematic analysis of qualitative data: AMEE Guide No. 131. Med Teach. 42 (8), 846–854. 10.1080/0142159X.2020.1755030

Killen, B.A., Brito da Luz, S., Lloyd, D.G., Carleton, A.D., Zhang, J., Besier, T.F., Saxby, D.J., 2021. Automated creation and tuning of personalised muscle paths for OpenSim musculoskeletal models of the knee joint. Biomech. Model. Mechanobiol. 20 (2), 521–533. 10.1007/s10237-020-01398-1

Lai, A.K.M., Arnold, A.S., Wakeling, J.M., 2017. Why are Antagonist Muscles Co-activated in My Simulation? A Musculoskeletal Model for Analysing Human Locomotor Tasks. Ann. Biomed. Eng. 45 (12), 2762–2774. 10.1007/s10439-017-1920-7

Lenaerts, G., Bartels, W., Gelaude, F., Mulier, M., Spaepen, A., Van der Perre, G., Jonkers, I., 2009a. Subject-specific hip geometry and hip joint centre location affects calculated contact forces at the hip during gait. J. Biomech. 42 (9), 1246–1251. 10.1016/j.jbiomech.2009.03.037

Lenaerts, G., De Groote, F., Demeulenaere, B., Mulier, M., Van der Perre, G., Spaepen, A., Jonkers, I., 2008. Subject-specific hip geometry affects predicted hip joint contact forces during gait. J. Biomech. 41 (6), 1243–1252. 10.1016/j.jbiomech.2008.01.014

Lenaerts, G., Mulier, M., Spaepen, A., Van der Perre, G., Jonkers, I., 2009b. Aberrant pelvis and hip kinematics impair hip loading before and after total hip replacement. Gait Posture. 30 (3), 296–302. 10.1016/j.gaitpost.2009.05.016

Liu, M.Q., Anderson, F.C., Schwartz, M.H., Delp, S.L., 2008. Muscle contributions to support and progression over a range of walking speeds. J. Biomech. 41 (15), 3243–3252. 10.1016/j.jbiomech.2008.07.031

Meyer, C.A.G., Wesseling, M., Corten, K., Nieuwenhuys, A., Monari, D., Simon, J.P., Jonkers, I., Desloovere, K., 2018. Hip movement pathomechanics of patients with hip osteoarthritis aim at reducing hip joint loading on the osteoarthritic side. Gait Posture. 59, 11–17. 10.1016/j.gaitpost.2017.09.020

Modenese, L., Barzan, M., Carty, C.P., 2021. Dependency of lower limb joint reaction forces on femoral version. Gait Posture 88, 318–321. 10.1016/j.gaitpost.2021.06.014

Moissenet, F., Modenese, L., Dumas, R., 2017. Alterations of musculoskeletal models for a more accurate estimation of lower limb joint contact forces during normal gait: A systematic review. J. Biomech. 63, 8–20. 10.1016/J.JBIOMECH.2017.08.025

Ng, K.C.G., Mantovani, G., Lamontagne, M., Labrosse, M.R., Beaulé, P.E., 2017. Increased Hip Stresses Resulting From a Cam Deformity and Decreased Femoral Neck-Shaft Angle During Level Walking. Clin. Orthop. Relat. Res. 475 (4), 998–1008. 10.1007/S11999-016-5038-2

Ng, K.C.G., Mantovani, G., Modenese, L., Beaulé, P.E., Lamontagne, M., 2018. Altered Walking and Muscle Patterns Reduce Hip Contact Forces in Individuals With Symptomatic Cam Femoroacetabular Impingement. Am. J. Sports Med. 46 (11), 2615–2623. 10.1177/0363546518787518

Passmore, E., Graham, H.K., Pandy, M.G., Sangeux, M., 2018. Hip- and patellofemoral-joint loading during gait are increased in children with idiopathic torsional deformities. Gait Posture 63, 228–235. 10.1016/j.gaitpost.2018.05.003

Rajagopal, A., Dembia, C.L., DeMers, M.S., Delp, D.D., Hicks, J.L., Delp, S.L., 2016. Full body musculoskeletal model for muscle-driven simulation of human gait. IEEE Trans. Biomed. Eng. 63 (10), 2068–79. 10.1109/TBME.2016.2586891

Samaan, M.A., Zhang, A.L., Popovic, T., Pedoia, V., Majumdar, S., Souza, R.B., 2019. Hip joint muscle forces during gait in patients with femoroacetabular impingement syndrome are associated with patient reported outcomes and cartilage composition. J. Biomech. 84, 138–146. 10.1016/j.jbiomech.2018.12.026

Savage, T.N., Saxby, D.J., Lloyd, D.G., Hoang, H.X., Suwarganda, E.K., Besier, T.F., Diamond, L.E., Eyles, J., Fary, C., Hall, M., Molnar, R., Murphy, N.J., O’donnell, J., Spiers, L., Tran, P., Wrigley, T.V., Bennell, K.L., Hunter, D.J., Pizzolato, C., 2022. Hip Contact Force Magnitude and Regional Loading Patterns Are Altered in Those with Femoroacetabular Impingement Syndrome. Med. Sci. Sports Exerc. 54 (11), 1831–41. 10.1249/MSS.0000000000002971

Scheys, L., Desloovere, K., Suetens, P., Jonkers, I., 2011. Level of subject-specific detail in musculoskeletal models affects hip moment arm length calculation during gait in pediatric subjects with increased femoral anteversion. J. Biomech. 44 (7), 1346–53. 10.1016/j.jbiomech.2011.01.001

Scheys, L., Van Campenhout, A., Spaepen, A., Suetens, P., Jonkers, I., 2008. Personalized MR-based musculoskeletal models compared to rescaled generic models in the presence of increased femoral anteversion: Effect on hip moment arm lengths. Gait Posture. 28 (3), 358–365. 10.1016/j.gaitpost.2008.05.002

Seth, A., Hicks, J.L., Uchida, T.K., Habib, A., Dembia, C.L., Dunne, J.J., Ong, C.F., DeMers, M.S., Rajagopal, A., Millard, M., Hamner, S.R., Arnold, E.M., Yong, J.R., Lakshmikanth, S.K., Sherman, M.A., Ku, J.P., Delp, S.L., 2018. OpenSim: Simulating musculoskeletal dynamics and neuromuscular control to study human and animal movement. PLoS Comput. Biol. 14 (7): e1006223. 10.1371/journal.pcbi.1006223

Shelburne, K., Decker, M., Krong, J., Torry, M., Philippon MJ, 2010. Muscle forces at the hip during squatting exercise. In: 56th Annual Meeting of the Orthopaedic Research Society; New Orleans, LA.

Shepherd, M.C., Clohisy, J.C., Nepple, J.J., Harris, M.D., 2023. Derotational femoral osteotomy locations and their influence on joint reaction forces in dysplastic hips. J. Orthop. Res. 10.1002/jor.25559

Shepherd, M.C., Gaffney, B.M.M., Song, K., Clohisy, J.C., Nepple, J.J., Harris, M.D., 2022. Femoral version deformities alter joint reaction forces in dysplastic hips during gait. J. Biomech. 135:111023. 10.1016/j.jbiomech.2022.111023

Song, K., Anderson, A.E., Weiss, J.A., Harris, M.D., 2019. Musculoskeletal models with generic and subject-specific geometry estimate different joint biomechanics in dysplastic hips. Comput. Methods Biomech. Biomed. Engin. 22 (3), 259–270. 10.1080/10255842.2018.1550577

Song, K., Gaffney, B.M.M., Shelburne, K.B., Pascual-Garrido, C., Clohisy, J.C., Harris, M.D., 2020. Dysplastic hip anatomy alters muscle moment arm lengths, lines of action, and contributions to joint reaction forces during gait. J. Biomech. 110:109968. 10.1016/j.jbiomech.2020.109968

Song, K., Pascual-Garrido, C., Clohisy, J.C., Harris, M.D., 2022. Elevated loading at the posterior acetabular edge of dysplastic hips during double-legged squat. J. Orthop. Res. 40 (9), 2147–55. 10.1002/jor.25249

Song, K., Pascual-Garrido, C., Clohisy, J.C., Harris, M.D., 2021. Acetabular Edge Loading During Gait Is Elevated by the Anatomical Deformities of Hip Dysplasia. Front. Sports Act. Living. 3:687419. 10.3389/fspor.2021.687419

Tateuchi, H., Yamagata, M., Asayama, A., Ichihashi, N., 2021. Influence of simulated hip muscle weakness on hip joint forces during deep squatting. J. Sports. Sci. 39 (20), 2289–97. 10.1080/02640414.2021.1929009

Thelen, D., Seth, A., Anderson, F.C., Delp, S.L., 2012. Gait 2392 and 2354 Models. OpenSim Documentation. Retrieved July 3, 2023, from: https://simtkconfluence.stanford.edu:8443/display/OpenSim/Gait+2392+and+2354+Models

Van Rossom, S., Emmerzaal, J., van der Straaten, R., Wesseling, M., Corten, K., Bellemans, J., Truijen, J., Malcorps, J., Timmermans, A., Vanwanseele, B., Jonkers, I., 2023. The biomechanical fingerprint of hip and knee osteoarthritis patients during activities of daily living. Clin. Biomech. 101:105858. 10.1016/j.clinbiomech.2022.105858

Vandekerckhove, I., Wesseling, M., Kainz, H., Desloovere, K., Jonkers, I., 2021. The effect of hip muscle weakness and femoral bony deformities on gait performance. Gait Posture. 83, 280–6. 10.1016/j.gaitpost.2020.10.022

Wesseling, M., Bosmans, L., Van Dijck, C., Vander Sloten, J., Wirix-Speetjens, R., Jonkers, I., 2019a. Non-rigid deformation to include subject-specific detail in musculoskeletal models of CP children with proximal femoral deformity and its effect on muscle and contact forces during gait. Comput. Methods Biomech. Biomed. Engin. 22 (4), 376–385. 10.1080/10255842.2018.1558216

Wesseling, M., Groote, F. De, Meyer, C., Corten, K., Simon, J.-P., Desloovere, K., Jonkers, I., 2016. Subject-specific musculoskeletal modelling in patients before and after total hip arthroplasty. Comput. Methods Biomech. Biomed. Engin. 19 (15), 1683–91. 10.1080/10255842.2016.1181174

Wesseling, M., Meyer, C., Corten, K., Desloovere, K., Jonkers, I., 2018. Longitudinal joint loading in patients before and up to one year after unilateral total hip arthroplasty. Gait Posture. 61, 117–124. 10.1016/J.GAITPOST.2018.01.002

Wesseling, M., Van Rossom, S., Jonkers, I., Henak, C.R., 2019b. Subject-specific geometry affects acetabular contact pressure during gait more than subject-specific loading patterns. Comput. Methods Biomech. Biomed. Engin. 22 (16), 1323–33. 10.1080/10255842.2019.1661393

Wu, T., Lohse, K.R., Dillen, L. Van, Song Phd, K., Clohisy, J.C., Harris, M.D., 2023. Are Abnormal Muscle Biomechanics and Patient-reported Outcomes Associated in Patients With Hip Dysplasia? Clin. Orthop. Relat. Res. 10.1097/CORR.0000000000002728

Yamaguchi G.T., Zajac F.E., 1989. A planar model of the knee joint to characterize the knee extensor mechanism. J. Biomech. 22 (1),1–10. 10.1016/0021-9290(89)90179-6. PMID: 2914967.

